# ClinVar and HGMD genomic variant classification accuracy has improved over time, as measured by implied disease burden

**DOI:** 10.1101/2022.10.26.22281567

**Authors:** Andrew G. Sharo, Yangyun Zou, Aashish N. Adhikari, Steven E. Brenner

## Abstract

Curated databases of genetic variants assist clinicians and researchers in interpreting genetic testing results. Yet these databases contain variants misclassified as pathogenic, which do not result in pathogenic phenotypes. Using archives of ClinVar and HGMD, we investigated how variant misclassification has changed over six years across different ancestry groups. We considered inborn errors of metabolism (IEMs) screened in newborns as a model system because these disorders are often highly penetrant with neonatal phenotypes. We used samples from the 1000 Genomes Project (1KGP) to identify individuals with genotypes that were classified by the databases as pathogenic. Due to the rarity of IEMs, nearly all such classified pathogenic genotypes indicate likely variant misclassification in ClinVar or HGMD. While the false positive rate of both ClinVar and HGMD have improved over time, HGMD variants currently would imply two orders of magnitude more affected individuals in 1KGP than ClinVar variants. We observed that African ancestry individuals have a significantly increased chance of being incorrectly indicated to be affected by a screened IEM when HGMD variants are used. However, this bias affecting genomes of African ancestry was no longer significant once common variants were removed in accordance with recent variant interpretation guidelines. We discovered that ClinVar variants classified as Pathogenic or Likely Pathogenic are reclassified 11-fold more often than DM or DM? variants in HGMD, which has likely resulted in ClinVar’s lower false positive rate. Considering misclassified variants that have since been reclassified, we found that variant interpretation guidelines and allele frequency databases comprised of genetically diverse samples are important factors in reclassification. Finally, we find that ClinVar variants common in European and South Asian individuals were more likely to be reclassified to a lower confidence category, perhaps due to an increased chance of these variants being classified by multiple submitters.

## Introduction

Rare genetic diseases may affect as many as 1 in 20 Americans^1^, but a definitive diagnosis is sometimes elusive^2^. In the past decade, exome and genome sequencing have improved the diagnostic rate for undiagnosed rare genetic diseases by 3 to 4-fold over previously established methods^2-4^. Identifying the causal variant(s) through sequencing can inform disease management by altering treatment, predicting disease progression, and informing risk to other family members including reproductive planning^5,6^. However, identifying causal variants can be challenging. Clinicians must objectively weigh many sources of evidence to determine if a variant explains the proband phenotypes. Indeed, the majority of individuals with a suspected rare genetic disease remain undiagnosed after exome or genome sequencing^2,7^.

To standardize the interpretation of variants, in 2015 the American College of Medical Genetics and Genomics (ACMG) and the Association for Molecular Pathology (AMP) developed guidelines to unify norms across clinical laboratories^8^. Since then, a growing number of laboratories have adopted these guidelines^9^. As familiarity with the guidelines has grown, variant interpretation concordance across laboratories has increased from 71% in 2016 to 84% in 2020^10,11^. These variant interpretation guidelines draw from several specialized research areas including population genetics, human gene isoforms, protein structure and function, and computational predictions of variant impact^12^. While these specialties have all made essential contributions to variant interpretation, perhaps no resource has been more valuable than the creation of diverse databases of allele frequencies, which are used to identify variants that are too common to cause a rare disease. In 2012, the Exome Sequencing Project created the first large-scale database of exonic allele frequencies that included samples from both European Americans and African Americans^13^. In 2015, phase 3 of the 1000 Genomes Project (1KGP) became available, providing genome-wide alleles from thousands of global genomes^14^. This was quickly followed by progressively larger and more diverse databases, including ExAC^15^, gnomAD^16^, and ALFA^17^. Here we investigate trends in the accuracy of variant interpretation since 2014, during which these allele frequency resources have grown tremendously.

Researchers communicate variant interpretations through published articles and submissions to variant databases. Until recently, variants were primarily classified in locus-specific databases (LSDBs) that typically collected variants in a single gene. In an effort to standardize content and improve ease of access, many LSDBs used the same software, the Leiden Open Variation Database^18^, and the Human Genome Variation Society collected LSDBs to form a databases of LSDBs^19^. Authoritative reference resources such as OMIM^20^ and GeneTests^21^ often included additional variant information. Following calls to harmonize these resources into a single common database^22^, today there are two leading genome-wide variant databases of clinical interest: ClinVar and the Human Gene Mutation Database (HGMD). HGMD began in 1996, is a commercial product with subscriptions sold by Qiagen, and is curated directly from published literature by dedicated staff. It contains pathogenic interpretations of nearly 300,000 variants. A free version of HGMD is available that is several years out of date. HGMD labels disease-causing variants as either disease-causing (DM) or likely disease causing (DM?). In 2013, the NIH created ClinVar, a free-to-access database (maintained by NCBI with input from ClinGen) that accepts submissions from clinical laboratories, research groups, and specialized databases. As of 2020, 8,000+ people access ClinVar each day, and it currently contains pathogenic interpretations of nearly 130,000 variants^23,24^. ClinVar labels disease-causing variants as either Pathogenic (P) or Likely Pathogenic (LP). By definition, P indicates a 99% chance of pathogenicity, and LP indicates a 90% chance of pathogenicity. Although these definitions provide an acceptable threshold for false positives, for the purposes of this paper, we highlight all variants that may be incorrectly classified as pathogenic.

ClinVar and HGMD attempt different strategies to reach the same goal: accurate variant classification. ClinVar receives variants primarily from clinicians and laboratory staff who often use standardized interpretation guidelines to identify pathogenic variants in a clinical context. In addition to cataloging pathogenic variants, ClinVar also catalogs variants classified as benign. HGMD curates information directly from research and clinical articles, which may include experimental assays of variant function^25^. These databases are rapidly growing. Since 2017, the number of ClinVar variants has doubled, and HGMD variants have grown by 50%.

Several studies have attempted to assess the accuracy of cataloged variants using large sequencing cohorts of healthy individuals^26-30^. Two of the earliest studies searched for variants classified as pathogenic in individuals sequenced in a population database created by the 1000 Genomes Project (1KGP)^14^. These researchers identified individuals in 1KGP who were homozygous for one or more recessive variants classified as pathogenic (henceforth, ‘indicated affected individuals’). Surprisingly, these two studies found that most individuals harbored multiple homozygous variants that were cataloged by HGMD to cause early-onset disease. However, individuals in 1KGP were all over 18 years of age and healthy enough to sign a consent form. Certainly, 1KGP individuals are not expected to be enriched for disease, yet these studies found that the implied rates of disease were higher in 1KGP than the known disease prevalence. There are two plausible explanations for this discrepancy. The first is that many benign variants were misclassified as pathogenic, which the authors concluded^28,29^. An alternative explanation is that some Mendelian diseases have been underdiagnosed. Since this is true for some disorders^31^, we chose to analyze a subset that are screened for at birth and are likely not substantially underdiagnosed (see Methods). With this modification, we believe that most, and likely all, of indicated affected individuals are not affected by a disease, and rather the classified pathogenic variants they harbor were misclassified (see caveats below). A similar approach has also been used to investigate ClinVar variants, which a 2018 study showed imply disease prevalence much higher than recorded prevalence for several clinically actionable or rare disorders^26^. Using orthogonal methods, researchers have identified variant features that are associated with correct classification. Specifically, they have found that recently curated variants, with lower minor allele frequency (MAF), with multiple concordant submissions, and submitted by clinical researchers are more likely to be correctly classified^30,32^.

Since many variants are found principally in a single ancestral population, misclassification can lead to disparities in variant interpretation and clinical care. Indeed, one study determined that variants erroneously associated with sudden heart failure were found at higher allele frequency in Black Americans than white Americans^33^. Fortunately, these misclassified variants were eventually corrected. However, until erroneously classified variants are corrected, which may take years, probands who harbor these variants may undergo inappropriate medical care. Furthermore, misclassified variants can have effects beyond the clinical care of individuals with those variants, since cataloged pathogenic variants can influence novel variant interpretation. In the ACMG/AMP variant interpretation guidelines, two categories of evidence that support pathogenicity rely directly on cataloged variants: the same amino acid change as a cataloged pathogenic variant (PS1) and a different amino acid change at the same residue as a cataloged pathogenic variant (PM5). Misclassified variants can also have indirect effects through the ACMG/AMP guidelines’ consideration of variant impact predictors, which contribute supporting evidence (PP3, BP4). Since many variant impact predictors are trained or are validated on cataloged variants ^34-37^, their predictions may be influenced by misclassified variants. In the worst case, a researcher following the ACMG/AMP guidelines may be misled by misclassified variants to incorrectly classify a novel variant, either by using misclassified variants as direct evidence (PS1, PM5) or indirectly though variant impact predictors that trained on misclassified variants (PP3, BP4). Such an event would propagate existing variant misclassifications and possibly reinforce disparities.

Variant databases have taken different approaches to address misclassifications. ClinVar introduced a star system to indicate the review status of a variant interpretation, in which a variant gains credibility when assertion criteria are provided, multiple submitters concur, or an interpretation comes from experts in the field who follow gene-specific classification guidelines^38^. Wright et al. found that variants classified as pathogenic with more review stars were more likely to be truly pathogenic^30^. ClinGen has also supported the formation of variant curation expert panels—composed of healthcare professionals with expertise relevant to a disease gene—which can provide high-confidence variant interpretations and resolve conflicting variant interpretations. Currently, ClinVar contains just 41 genes in which 10 or more variants are reported as reviewed by an expert panel, out of more than 3,000 genes associated with a monogenic disorder by OMIM^20^. Although expert panels are promising, they have so far contributed to a small fraction of ClinVar variant reclassifications. HGMD curators reclassify variants based on new published evidence such as functional studies or population frequency, and their reclassification rate has been reported as similar to that of ClinVar^25,39^. Here, we consider whether these reclassification efforts, in concert with improved resources, have reduced the number of apparently misclassified variants over time. We consider variants in a subset of well-studied genes with highly penetrant phenotypes.

Inborn errors of metabolism (IEMs) are a group of rare, primarily recessive or X-linked, monogenic disorders caused by defects in a metabolic enzyme or its cofactors. Newborns in most developed countries are screened for IEMs using blood metabolites. Untreated, many of these screened IEMs are highly penetrant and lead to metabolite accumulation that often causes irreversible disability or death. They are thus a model system for identifying false positives in variant databases, as they should not be present as pathogenic genotypes in healthy individuals. While many screened IEMs are debilitating or fatal in childhood unless treated, there are notable exceptions. For example, our screened IEMs include Short Chain Acyl CoA Dehydrogenase Deficiency (SCADD; associated with *ACADS*) and Hyperprolinemia type I (HPI; associated with *PRODH*), both of which often don’t yield disease^40,41^. Additionally, our screened IEMs include Ornithine transcarbamylase deficiency (OTCD; associated with *OTC*) and Glutaric Acidemia Type II (GAII; primarily associated with *ETFDH*), both of which are often seen in late-onset forms which may not result in outward symptoms until adulthood^42,43^.

Because screened IEMs are systematically identified in the population, their maximum possible incidence is generally known, and there has been greater opportunity to identify and catalog the genetic variants that cause these diseases. Indeed, one recent study found potential benefit to screening newborns for IEMs using exome sequencing alongside mass spectrometry, the current standard for screening^44^. However, these researchers found it necessary to manually curate dozens of variants cataloged in ClinVar or HGMD for which the MAF was higher than expected for a rare disorder. Out of 60 variants with MAF > 0.1%, they deemed 41 were not reportable due to insufficient published evidence for pathogenicity.

Variants with a MAF greater than expected from disease incidence are addressed in the 2015 ACMG/AMP variant interpretation guidelines^8^ under the BA1 evidence for benign variants. These guidelines recommend that a MAF >5% in 1KGP, ExAC (now superseded by gnomAD), or the Exome Sequencing Project (ESP) may be considered stand-alone evidence that the variant is benign. In 2018, the guidelines for this classification were updated by Ghosh et al. to recommend that a MAF >5% in any continental population dataset of at least 2,000 alleles (with some additional constraints) is stand-alone evidence the variant is benign^45^. We have investigated how implementing the original vs revised guidelines impacts our results.

Here, we investigate how the degree of variant misclassification has changed over time in ClinVar and HGMD, using screened IEMs as a model system. Building on previously developed methods^28,29^, we used samples in the 1000 Genomes Project (1KGP) to identify individuals who harbor genetic variants that have been listed in ClinVar or HGMD as pathogenic. We identified more individuals than expected from screened IEM incidence, an indication of the specificity of each database. We investigated how the number of these likely false positive individuals indicated by ClinVar and HGMD changed over time, and we considered whether certain ancestry groups were over-represented. Since we do not measure false negatives, we cannot assess the sensitivity of each database even though the balance between specificity and sensitivity is an important tradeoff to consider. We looked in detail at variants that were misclassified and what led to their eventual reclassification. Additionally, we probed overall trends of reclassification in ClinVar and HGMD, identifying surprising trends in the reclassification of confidently classified variants to uncertain classifications. Finally, we replicated our findings using samples from gnomAD, which includes 63,269 genomes.

## Materials and Methods

### Identifying indicated affected individuals in 1KGP

We used GRCh38 genotypes from 1KGP phase 3^14^ VCF files (downloaded on 14 November, 2019) to identify individuals who harbor genotypes classified as pathogenic (defined as homozygous, hemizygous, or compound heterozygous) but who likely do not suffer from a screened IEM. Ancestry was determined by superpopulation membership, as listed by the International Genome Sample Resource^46^. We created a curated list of 80 genes (Table S2), associated with 48 IEMs screened by the California newborn screening program^47^ (henceforth, screened IEMs). These screened IEMs include some disorders where a large fraction of affected individuals is asymptomatic. In our analysis below, we identified several ClinVar variants in *PRODH*, associated with HPI. This condition is characterized by elevated levels of proline, and it is sometimes considered benign and asymptomatic^41^. However, there are reports of individuals with HPI who have severe neurological impairment^48^. Additionally, recent long-term follow-up of patients with HPI suggests it results in impaired social skills, and there is evidence that deletions containing *PRODH* (and possibly variants in *PRODH*) contribute to schizophrenia risk^49-51^. Given the possible clinical phenotypes associated with this gene, we retained it in our analysis.

The population incidence of screened IEMs is approximately 1 in 3,200^52^. Thus, if the individuals sequenced in 1KGP were a random sample with unknown health status at birth, we would expect less than 1 individual to have a screened IEM. Given that most of the indicated affected individuals lived in countries without newborn screening programs before 1990 (Table S1), they are unlikely to have been screened and treated early enough to prevent irreversible damage.

ClinVar GRCh38 variants were obtained from VCF files (downloaded on 8 January, 2021) from the NCBI ClinVar FTP site^38^. VCF files were gathered from both archives 1.0 and 2.0 (starting with clinvar_20140401.vcf.gz and ending with clinvar_20201226.vcf.gz). Bcftools norm^53^ was used to left-align and normalize indels. Only variants within our list of 80 genes were considered further. Variants that were listed as only somatic or variants with null alt alleles were not considered further. For ClinVar archive 1.0 variants, variants were assigned clinical significance using the following categories: ‘0’: VUS, ‘2’: Benign (B), ‘3’: Likely benign (LB), ‘4’: Likely pathogenic (LP), and ‘5’: Pathogenic (P). Variants were inferred to have Conflicting interpretations when they had interpretations in two or more of the following three categories: B or LB, VUS, P or LP. For ClinVar archive 2.0 variants, variants were assigned clinical significance using the following categories: ‘Benign’: B, ‘Benign/Likely_benign’: B, ‘Likely_benign’: LB, ‘Uncertain_significance’: VUS, ‘Likely_pathogenic’: LP, ‘Pathogenic/Likely_pathogenic’: P, ‘Pathogenic’: P, ‘Conflicting_interpretations_of_pathogenicity’: Conflicting. For variants with multiple interpretations separated by commas, if one of the interpretations was in the above list of categories, the variant was assigned to that category (e.g., ‘Pathogenic,_risk_factor’ would be assigned to P). Due to inconsistencies in review star annotation in archive 1.0 files before June 15, 2015, ‘not’ was assigned 0 review stars, ‘single’ was assigned as 0or1 review stars (see below for details), ‘conf’ was assigned as 1 review star, and ‘mult’ was assigned as 2 review stars. For archive 1.0 files after June 15, 2015, ‘no_assertion_criteria_provided’, ‘no_assertion_provided’, ‘not’, ‘no_criteria’, and ‘no_assertion’ were grouped as 0 review stars; ‘criteria_provided’, ‘conf’, and ‘single’ were grouped as 1 review star; ‘_multiple_submitters’, ‘_no_conflicts’, and ‘mult’ were grouped as 2 review stars. For all archive 1.0 files, review stars were assessed manually for variants with an inferred pathogenic genotype in 1KGP. For archive 2.0 variants, “no_assertion_criteria_provided”, “No_assertion_provided”, and “no_interpretation_for_the_single_variant” were grouped as 0 review stars, “criteria_provided”, “_single_submitter”, and “_conflicting_interpretations” as 1 review star, “_multiple_submitters”, “_no_conflicts”, and “reviewed_by_expert_panel” as 2+ review stars. In calculating indicated affected individuals for each year, we reported the maximum number of individuals with an inferred pathogenic genotype at any time in that year. In our analysis of 1KGP indicated affected individuals, ClinVar submissions were removed from consideration if the submitted condition was a not a screened IEM (e.g., Schizophrenia). Submissions for which the condition was “not provided” were included in our analysis. For all other analyses, it was not feasible to check the submitted condition of variants.

HGMD variants were obtained from privately archived versions of HGMD 2014.1 and 2016.2, and a recently accessed version of 2020.3 through Qiagen Digital Insights HGMD Professional. Only SNVs classified at least once as ‘DM’ or ‘DM?’ within our list of 80 screened IEM genes were considered further. There were a handful of variants with multiple classifications, and these were assigned the most severe classification.

In our analysis using the 2015 BA1 guidelines, variants with a global MAF > 5% in 1KGP, the Exome Sequencing Project (ESP6500SI-V2), or gnomAD v2.1 exomes were removed from consideration. In our analysis using the 2018 BA1 guidelines, variants with a global MAF > 5% in 1KGP or ESP, or a MAF >5% in any gnomAD exome continental population were removed.

Ensembl Variant Effect Predictor with custom annotations was used to annotate the 1KGP VCF with all features. For rapid I/O of VCFs, we used cyvcf2^54^. To identify when the ancestry composition of indicated affected individuals (aggregated across all screened IEMs) was significantly different from the ancestry composition of 1KGP or gnomAD, we first performed a two-sided Fisher’s exact test on a 5 × 2 contingency table that included the five continental populations (African, Latino, East Asian, European, South Asian), using fisher.test in the R ‘stats’ package^55^. When the expected count for every population was greater than 40, we instead performed a Pearson’s Chi-squared test using chisq.test to reduce computation time. For those global analyses that showed significant deviation from the 1KGP database ancestry composition, we performed individual tests to identify the significantly skewed population. These individual tests were performed using a one-sided Fisher’s exact test on a 2 × 2 contingency table as described above. To correct for multiple tests, we used a 5% significance threshold with Bonferroni correction for 222 tests, yielding a p-value threshold of 2.2 × 10^−4^. We determined 222 tests by calculating the total number of tests performed across all figures (including supplementary figures), which were typically 1 Fisher’s exact test per bar, with an additional 5 tests per bar when the Fisher’s exact test was significant. Bars that had zero height were not tested. Odds ratios and 95% confidence intervals were determined using two-sided Fisher’s exact tests as described above.

To confirm that the inferred pathogenic genotypes we observed in 1KGP were not sequencing errors, we attempted to confirm the quality of all variants that comprised these genotypes (Table S3). Specifically, we downloaded whole genome and deep exome sequencing BAM alignment files of select individuals with homozygous, hemizygous, or compound heterozygous inferred pathogenic genotypes. Most of these alignments were improved by quality-control steps including marking duplicates, local realignment around indels, and base quality recalibration, especially for the Illumina sequencing data. Next, we detected variants and calculated genotypes for each sample at specific sites based on both low-coverage genome sequencing data (<5×per site per individual) and high-coverage exome sequencing information (at least >20×per site per individual) using ‘UnifiedGenotyper’ from the Genome Analysis Toolkit (GATK 3.4-0) under a multi-sample calling strategy^56,57^. Variant Quality Score Recalibration (VQSR) was conducted to evaluate variant quality by GATK 3.4-0. Finally, we obtained variant and genotype information of select individuals and their site-specific genotype quality parameters such as genotype quality (GQ) to validate the quality of the called genotypes. We used GQ≥30 (p-value of 0.001) as our threshold for high quality genotype calls. Genotypes of some individuals (Table S3) were re-confirmed based on high-coverage whole genome sequencing by Complete Genomics^58^. Thanks to the recent availability of high-coverage whole genome sequencing of all 1KGP samples from the New York Genome Center^59^, the remaining inferred pathogenic genotypes were confirmed using these data. Two inferred pathogenic genotypes in PRODH were not able to be reconfirmed due to poor sequencing quality in the gene.

To infer screened IEM incidence from 1KGP, for each IEM gene *g*, we summed the allele frequencies of all classified pathogenic variants in *g*, which we call *p*_*g*_. Genes were then divided into two categories: X chromosome and autosomal. The disease incidence for all X-linked disorders was calculated as 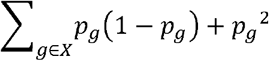 where *X* is the set of all X-linked screened IEM genes. For all autosomal genes, the incidence was calculated as 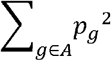 where *A* is the set of all autosomal screened IEM genes. We repeated this process for each population using the 1KGP population-specific MAF as well. In Figs. S3-5, the height of each bar represents the incidence inferred using the database-wide MAF, while the proportion of the bar comprised by each ancestry is based on the relative disease incidence calculated using the population-specific MAF. The same process was repeated for our gnomAD analysis in Figs. S8-10

### Variant reclassification in ClinVar and HGMD

ClinVar and HGMD variants were filtered as described above. VEP^60^ was used to annotate each variant with its gnomAD v2.1 exomes MAF. In order to identify reclassifications, for each time point available for ClinVar, each variant was classified into one of the following categories: B/LB 3 stars, B/LB 2 stars, B/LB 1 star, B/LB 0 stars, VUS, Conflicting, P/LP 0 stars, P/LP 1 star, P/LP 2 stars, P/LP 3 stars. At each time point available for HGMD, each variant was classified into one of the following categories: DM, DM?, DFP, DP, R. HGMD variants that were removed from the database were classified as R. Variants classified in any other category (such as ‘not provided’) and all ClinVar variants prior to June 15, 2015 were not considered. To create figures for the Results section ‘Comparison of variant reclassification between ClinVar and HGMD’, for each variant we considered only its first category chronologically (typically its category when first entered into the database) and its last category chronologically.

Next, for each ClinVar variant we used gnomAD v2.1 exomes to determine the ancestry group in which it occurs at the highest MAF. To reduce bias from the unequal number of individuals in each ancestry group in gnomAD, all ancestry-specific MAFs below 6.152×10^−5^ (the smallest possible MAF in African ancestry, which has the smallest number of individuals in gnomAD) were set to zero. Next, each variant was assigned to the ancestry with the highest MAF. Variants with zero MAF in all ancestries were not considered further.

For each variant, we recorded all reclassifications it underwent. To avoid classifications without stars, only ClinVar reclassifications after June 15, 2015 were considered. ClinVar GRCh38 VCF files (as described above) were used to identify reclassifications. Reclassifications were considered every month. Since more recent ClinVar VCFs were archived weekly, these were downsampled to approximate monthly archives. The removal of a ClinVar variant from the database was not considered a reclassification. If a variant re-entered into ClinVar under a new classification, it was considered reclassified.

Variant reclassifications were grouped into two categories: increasing confidence and decreasing confidence. Increasing confidence was defined as Conflicting or VUS to P/LP or B/LB with any number of stars. Decreasing confidence was defined as: P/LP or B/LB with any number of stars to Conflicting or VUS. Variants were grouped by these categories, colored by assigned ancestry (see above), and visualized using Floweaver^61^, resulting in Fig. 2c,e.

**Figure 1.**
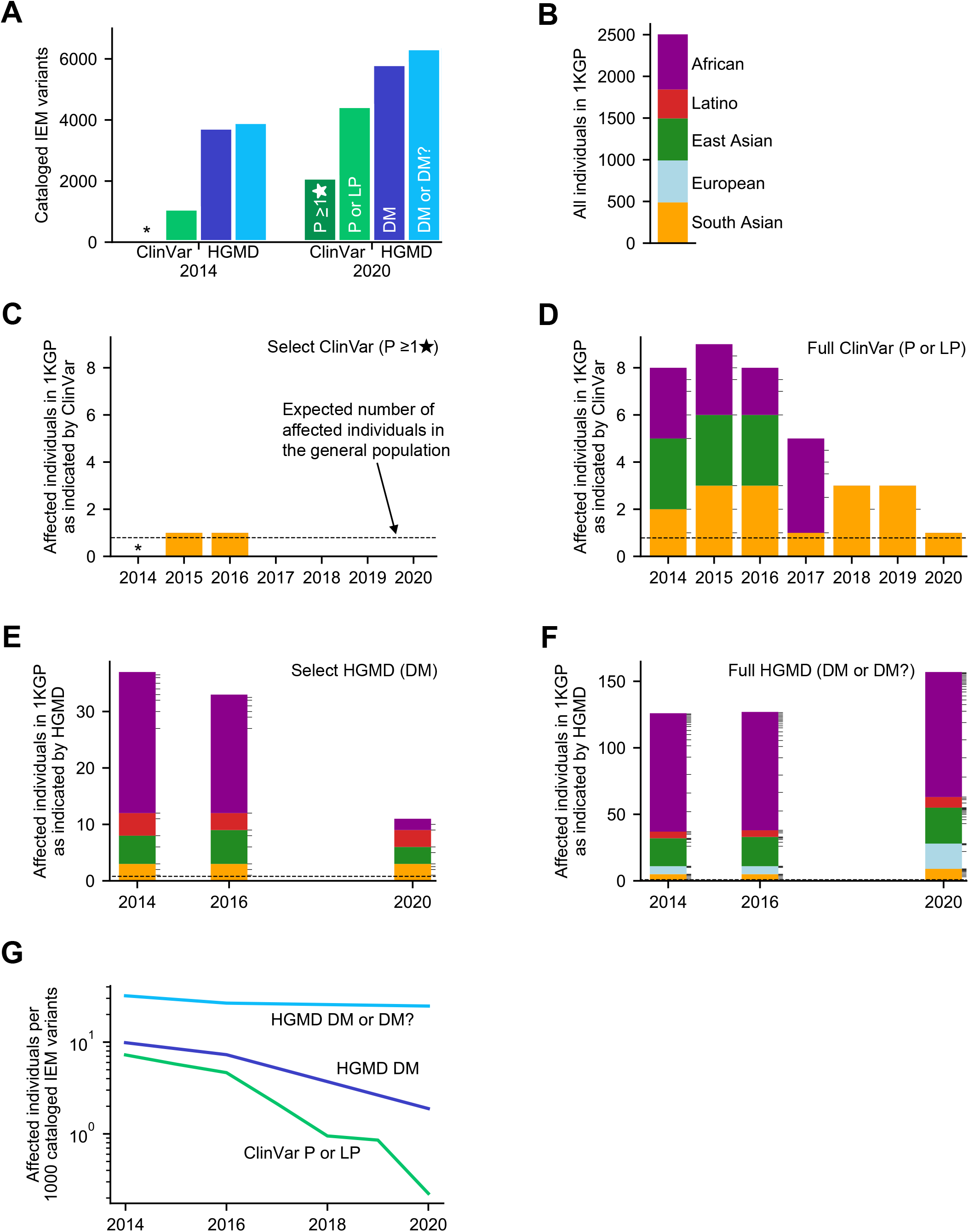
Number of 1KGP individuals indicated affected for screened IEMs by ClinVar or HGMD over time. (A) Number of screened IEM variants present in ClinVar or HGMD in 2014 and 2020. (B) Ancestry composition of individuals in 1KGP. (C-F) Bars are colored by ancestry as shown in (B). Tick marks on bars cluster individuals by the variant classified as pathogenic that they harbor. Dashed black lines indicate the aggregate population incidence of screened IEMs. The number of 1KGP individuals with an implied pathogenic genotype for a variant in (C) Select ClinVar variants classified as pathogenic, defined as variants with a P interpretation with at least 1 review star. Variants that also have conflicting interpretations (with VUS or B/LB) with 1 or more review stars are removed. (D) Full ClinVar variants classified as pathogenic, defined as variants with a P or LP interpretation. Variants that also have conflicting interpretations (with VUS or B/LB) are removed. (E) Select HGMD variants, defined as variants classified as DM. 2014, 2016, and 2020 are shown because they are the years for which we have archived HGMD data. (F) Full HGMD variants, defined as variants classified as DM or DM?. (G) The number of affected individuals relative to the number of variants classified in each variant set. This approximates a false positive rate, which has fallen over time for each database. *Data not available because existing review star framework was not in place until 2015.

**Figure 2.**
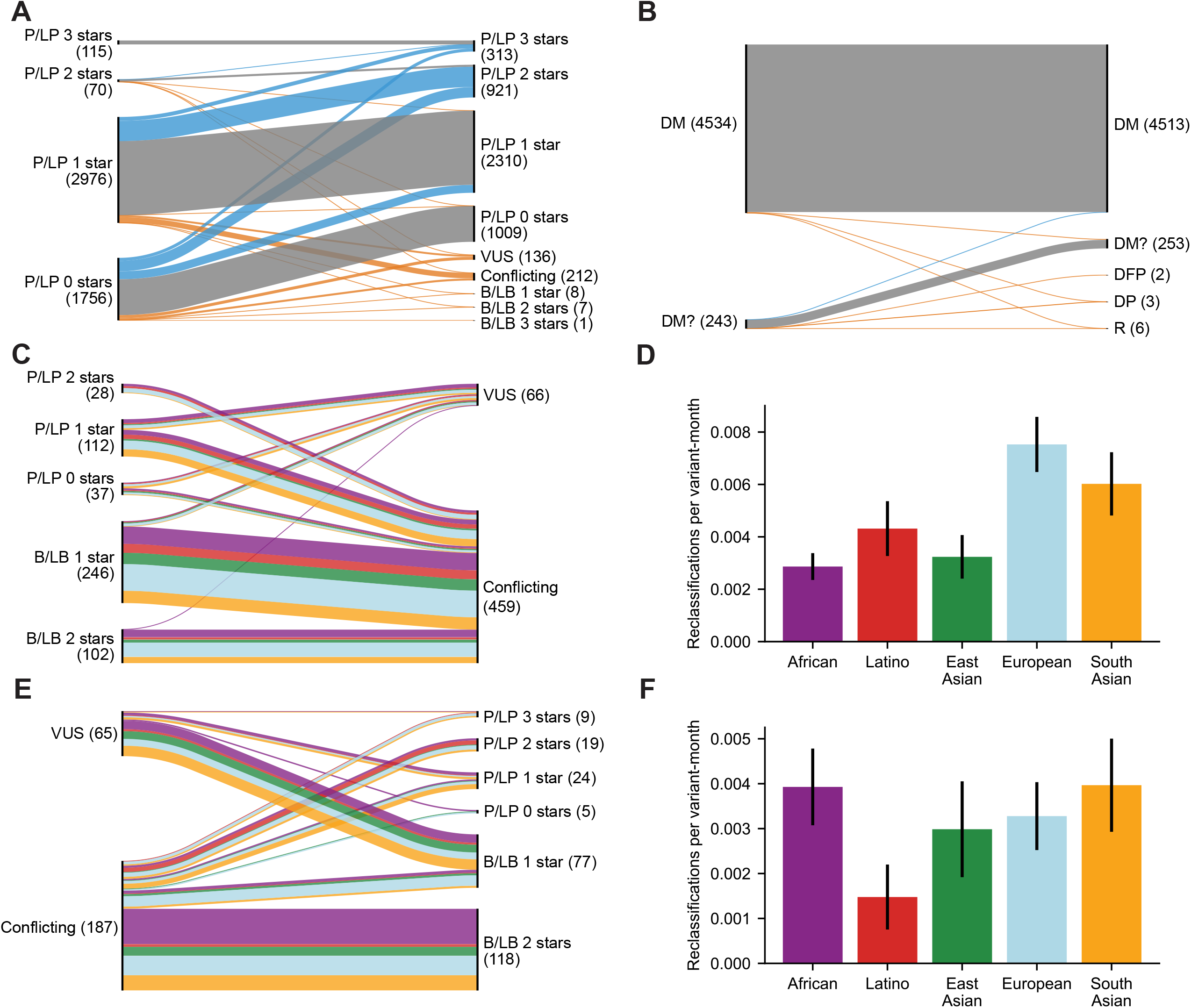
Variant reclassification in ClinVar and HGMD. (A) Reclassification paths of P/LP screened IEM ClinVar variants from 2014 (or first submission thereafter) to 2020, visualized in a Sankey plot in which line width represents the number of reclassified variants. Blue lines indicate increasing pathogenicity or review stars, orange lines indicate increasing benignity or reduced confidence of pathogenicity, and gray lines indicate no change. Numbers in parentheses provide variant counts of initial and final classifications for each category. (B) Reclassification paths of DM and DM? HGMD variants from 2014 to 2020. Disease-associated polymorphism (DP) and disease-associated polymorphism with additional functional evidence (DFP) are used to classify variants associated with disease but not necessarily disease-causing. Variants are retired (R) when they are found to no longer be associated with disease. (C) Reclassification paths of ClinVar variants from P/LP or B/LB to VUS or Conflicting. We plot only variants that could be assigned to a principal ancestry. Variant paths are colored by ancestry as in (D). (D) Rate of reclassification of variants shown in (C) when normalized by the historical ancestry composition of variants in ClinVar. (E) Reclassification paths of ClinVar variants from VUS or Conflicting to P/LP or B/LB. (F) Rate of reclassification of variants shown in (E) when normalized by historical ancestry composition of variants in ClinVar.

To correct for bias caused by the possible overrepresentation of some ancestries in ClinVar, for each ancestry we calculated the number of variants in each classification category. The number of variants per category was were calculated for every month, yielding a measure we call variant-months. A variant-month is a measure of both the number of variants and how long they have been in ClinVar. For example, 2 variants classified in ClinVar for a month is 2 variant-months, and 1 variant classified in ClinVar for 2 months is also 2 variant-months. For each ancestry, we analyzed its assigned variants to determine how many variant-months were cataloged for each category between June 15, 2015 and Dec 31, 2020. The differences in variant-months between ancestries reflects differences in genetic diversity as well as ClinVar submission bias. These variant-months are used to normalize comparisons across ancestries which we report in reclassifications per variant-month. In normalizing a reclassification category (increasing confidence or decreasing confidence), we divide the number of reclassifications by the variant-months of the source category. For example, if we wanted to compare increasing confidence across ancestries, then for each ancestry we would calculate the number of reclassifications with increasing confidence among variants assigned to that ancestry and divide that by the variant-months of the source category, in this case VUS and Conflicting variants. 95% confidence intervals were calculated for each ancestry group as +/- 1.96*sqrt(p*(1-p)/n) where p is reclassified variants / variant-months of source variants and n is variant-months of source variants. To identify significant reclassification rate differences between ancestries, we first performed a two-sided Fisher’s exact test on a 5 × 2 contingency table (as described above) containing the number of reclassifications and number of variant-months of the source category for each ancestry. We then performed one-sided Fisher’s exact tests on 2 × 2 tables (as described above) for each possible pair of ancestries.

## Results

### Individuals indicated as affected by ClinVar variants

We analyzed ClinVar screened IEM variants submitted between April 2014 and December 2020, and first examined a Select subset based on review stars (see Methods). This Select subset included P variants with 1 or more review stars (indicating the submitter included assertion criteria), which consisted of 2,118 variants in 2020 (Fig. 1A). In accordance with the 2015 ACMG/AMP BA1 guidelines, we removed variants with a MAF that reached the threshold for classification as stand-alone benign (global MAF > 5% in 1KGP, gnomAD, or ESP). This resulted in the removal of a single variant with 1 review star and a global MAF of ∼5% (Table S4). We later discuss applying the 2018 BA1 guidelines. To identify individuals who harbored inferred pathogenic genotypes of these Select ClinVar variants, we used the 1KGP database. 1KGP includes 2,504 individuals that are drawn approximately evenly from 5 continental populations (Fig. 1B). We considered individuals to be indicated affected if they were homozygous, hemizygous, or compound heterozygous for one or more Select variants. We found a single indicated affected individual, with South Asian ancestry, who was homozygous for a P variant (ACADS:c.1108A>G) added to ClinVar in 2015, which was re-classified as Conflicting by 2017 (Fig. 1C; Table 1). There have since been zero indicated affected individuals through 2020.

**Table 1:** Subset of ClinVar variants seen in a pathogenic genotype in 1KGP. See Table S6 for full list of ClinVar variants. NC indicates no assertion criteria were provided by the submitter.

| Chr | Position | Ref | Alt | Gene | cDNA,<br>protein | # hom<br>or hemi<br>in 1KGP | # comp<br>het in<br>1KGP | 1KGP<br>sample<br>ID(s) | First pathogenic submission:<br>submitter, date, interpretation,<br>evidence | Consensus<br>interpretation as of<br>Dec 2020 | Submitted<br>Interpretations |
| --- | --- | --- | --- | --- | --- | --- | --- | --- | --- | --- | --- |
| 9 | 130458549 | G | T | ASS1 | c.323G>T<br>R108L | 1 | 1 | NA19030<br>NA19395 | OMIM,<br>April 2014,<br>Pathogenic NC,<br>Heterozygous variant in affected<br>individual | Conflicting<br>Interpretations of<br>Pathogenicity | 1 Pathogenic NC<br>1 VUS<br>4 Likely benign<br>1 Benign<br>1 Benign NC |
| 12 | 109561798 | C | T | MMAB | c.403G>A<br>p.A135T | 1 | 0 | HG03169 | GeneReviews,<br>February 2016,<br>Pathogenic NC,<br>Seen in affected individuals | Conflicting<br>Interpretations of<br>Pathogenicity | 1 Pathogenic NC<br>1 VUS<br>1 Benign<br>1 Benign NC |
| 12 | 120739317 | A | G | ACADS | c.1108A>G<br>p.M370V | 1 | 0 | NA20878 | GeneDx,<br>August 2015,<br>Pathogenic,<br>clinical testing | Conflicting<br>Interpretations of<br>Pathogenicity | 2 VUS<br>1 Likely Benign |
| X | 38367361 | G | A | OTC | c.148G>A<br>p.G50R | 1 | 0 | NA21124 | GenMed Metabolism Lab,<br>April 2014,<br>Pathogenic NC,<br>Identified in late onset individual | Pathogenic, 0 stars | 2 Pathogenic NC |

In addition to Select ClinVar variants, clinicians often consider and report P and LP variants with 0 review stars (no assertion criteria) but give them appropriately lower credence. To analyze these variants, we next considered the Full dataset of ClinVar screened IEM variants, which included P and LP variants with any number of review stars. We removed from consideration variants that fulfilled the 2015 BA1 criteria. This eliminated six variants from 2014 to 2020, with a median MAF in 1KGP of 12% (Table S4). We searched for individuals in 1KGP who were indicated affected (Fig. 1D). In 2014, there were 8 indicated affected individuals, which increased to 9 in 2015, and declined to just 1 by 2020 (reclassification causes discussed below). 11 variants played a role in the genotypes of these indicated affected individuals (Table S6). We also considered whether P or LP variants led to a larger number of indicated affected individuals. However, due to the relatively small fraction of variants that are classified as LP, the results of considering only P variants were nearly identical to considering both P and LP (Fig. S1).

We did not observe any statistically significant skew in the ancestries of the 1KGP individuals who were indicated affected.

### Individuals indicated as affected by HGMD variants

Similar to our ClinVar analysis, we first examined a Select subset of HGMD variants. This subset included HGMD DM (disease-causing) variants in any screened IEM gene, which consisted of 5,833 variants in 2020. We removed one variant that met the 2015 BA1 criteria in 2014 and 2016 with a MAF of 20%. We removed an additional variant in 2016 with a MAF of 50% (Table S4). We investigated individuals in 1KGP who harbored Select HGMD variants, and we found 37 indicated affected individuals in 2014, caused by 16 variants (Fig. 1E). Repeating this analysis with Select HGMD classifications from December 2020, we found 11 indicated affected individuals in 1KGP (70% reduction from 2014) due to 9 variants (reclassification causes discussed below). 3 of these 9 variants were added to HGMD after 2014.

To gain a larger picture of potential variant misclassification in HGMD, we next considered the Full dataset of HGMD variants classified to likely cause disease, which included DM and DM? variants (henceforth, Full HGMD variants). We removed 5 variants that met the 2015 BA1 criteria in 2014, 4 variants in 2016, and 7 variants in 2020 (Table S4). We investigated individuals in 1KGP who harbored Full HGMD variants. In 2014 there were 126 indicated affected individuals in 1KGP due to 20 DM and 12 DM? variants (Fig. 1F). This increase in the number of DM variants compared to our Select analysis is due entirely to compound heterozygotes consisting of one DM variant and one DM? variant. Unexpectedly, we found indicated affected individuals increased over time, with 157 individuals in 2020, due to 17 DM and 27 DM? variants. These include 7 DM? and 4 DM variants that were added to HGMD since 2014.

These indicated affected individuals are not only a barometer for changes in potentially misclassified variants, but they can also inform whether particular ancestry groups are more likely to be affected by variant misclassifications. Considering Select HGMD variants in 2014, African ancestry individuals were significantly more likely to be indicated affected (Fig. 1E). While 26.4% of individuals in 1KGP are of African ancestry (Fig. 1B), 25 out of 37 (67.6%) of indicated affected individuals had African ancestry, which is significantly more than expected by chance (p < 10^−6^) and indicates an odds ratio of 5.8 for African ancestry individuals (95% CI: 2.8-12.8). By 2020, no populations were significantly skewed. Notably, in 2014, 2016, and 2020, no European ancestry individuals were indicated affected. When considering Full HGMD variants, we found that 89 out of 126 indicated affected individuals in 2014 and 94 out of 157 indicated affected individuals in 2020 were of African ancestry (both p < 10^−15^) (Fig. 1F). This translates to an odds ratio of 6.7 (95% CI: 4.5-10.2) in 2014 and 4.2 (95% CI: 3.0-5.9) in 2020. Unlike the ancestry skew observed in Select HGMD variants, the ancestry skew in Full HGMD variants has persisted over time.

### False positive rate across datasets

Since each ClinVar or HGMD dataset contains a different number of cataloged IEM-associated variants (Fig. 1A), we developed a metric to enable a comparison of false positive rate across datasets. For each available year, we calculated the number of indicated affected individuals in 1KGP divided by the number of cataloged variants. Although we cannot be certain that no individual in 1KGP has a screened IEM, this metric is a proxy for the false positive rate of each database. In 2014, the Full ClinVar dataset indicated 7.3 affected individuals per 1,000 cataloged P or LP variants (Fig. 1G). By 2020, this false positive rate had decreased by 97%. We could not determine a meaningful false positive rate for the Select ClinVar dataset due to the several years with zero indicated affected individuals. For Select HGMD variants, the false positive rate decreased by 81%, with most of this decrease occurring between 2016 and 2020 (Fig. 1G). For Full HGMD variants, the false positive rate decreased 26% from 2014 to 2020 (Fig. 1G). It may seem surprising that the false positive rate of Full HGMD variants is decreasing given the increase in indicated affected individuals over time (Fig. 1F). However, this decrease is due to the ∼60% growth in cataloged variants between 2014 and 2020, which outweighed the growth in indicated affected individuals.

These three datasets have reduced the false positive rate of their cataloged variants over time, yet false positive rates currently differ greatly between them. As of 2020, Full ClinVar variants indicate 0.22 affected individuals per 1,000 cataloged pathogenic variants, which is an order of magnitude lower than Select HGMD variants, which indicate 1.9 affected individuals per 1,000 cataloged pathogenic variants (Fig. 1G). This, in turn, is an order of magnitude lower than Full HGMD variants, which indicate 25 affected individuals per 1,000 cataloged pathogenic variants.

### Reliability of genotypes

To ensure that the inferred pathogenic genotypes we observed in 1KGP were not caused by errors from sequencing or downstream variant and genotype calling, we independently confirmed nearly all Select ClinVar variants, Full ClinVar variants, and Select HGMD variants present in an inferred pathogenic genotype (Table S3). We re-called a subset of these genotypes using available low-coverage genome sequencing and high-coverage exome sequencing data from 1KGP (see Methods). We found that nearly all variants classified as pathogenic in this subset passed variant quality score recalibration (VQSR) filtering, and most genotypes in the indicated affected individuals had a genotype quality (GQ) larger than 30. For variants that we did not attempt to re-call or for which re-call quality was poor, we confirmed genotypes using high-coverage whole genome sequencing by either Complete Genomics or the New York Genome Center (see Methods). Out of the entire set of 52 genotypes indicated as pathogenic, there were just two for which genotype quality was below 30. One was TAZ:c.383T>C present in Select HGMD variants and found in a hemizygous state in HG03196. The other genotype consisted of a pair of compound heterozygous variants (PRODH:c.1357C>T;c.1322T>C) present in Full ClinVar variants and harbored by NA19372. Overall, we confirmed that 96% of the inferred pathogenic genotypes are high quality and reproducible. This suggests that the over-representation of putatively pathogenic genotypes in 1KGP is unlikely to be explained by errors introduced by sequencing or data processing.

### ClinVar variant reclassification

Between 2014 and 2020, 11 variants in the Full ClinVar dataset were part of an inferred pathogenic genotype in at least one 1KGP individual (Table S6). As of December 2020, 10 of these 11 variants have been reclassified in ClinVar to a non-pathogenic category. 8 variants were reclassified to Conflicting, 1 variant to VUS, and 1 variant to B/LB. One variant remains classified as P with 0 review stars. These variants were present in 7 genes: *OTC* (3), *ASS1* (2), *PRODH* (2), *ACADS* (1), *MMAB* (1), *MMUT* (1), and *SLC22A5* (1). Variants within the same gene tended to be initially contributed by the same submitter. For example, GenMed Metabolism Lab submitted the first interpretation for all three variants in *OTC*, and OMIM first provided both *PRODH* variants. For each variant, we also recorded the submitter that contributed the first non-pathogenic classification but did not identify any patterns (Table S6).

Among these 11 variants, we noticed a trend in which variants were initially submitted as P or LP when seen in an affected individual, even though there was limited evidence for pathogenicity. As more information because available, such as MAF, later submitters, most using defined criteria, interpreted these variants as VUS, B, or LB. One illustrative case is the variant A135T in *MMAB* (Table 1). Through a semi-automated process, this variant was extracted from a GeneReviews table to a ClinVar record in February 2016 as P and included two articles to support the interpretation^62,63^. According to these articles, researchers found this variant in a heterozygous state in three African ancestry individuals with methylmalonic acidemia (MMA) cblB type. In addition to A135T, each of these individuals also harbored a suspected pathogenic variant, although it was not confirmed to be in trans. Both articles claim the A135T variant was absent from control samples, for which ancestry information was not provided. We now know the MAF of this variant in African ancestry individuals is approximately 1% in 1KGP and gnomAD exomes, corresponding to a disease incidence of 1 in 10,000 assuming complete penetrance. However, MMA cblB type occurs in less than 1 in 50,000 births, and has not been seen at elevated levels in individuals of African ancestry^52^. This variant was observed in a homozygous state in an African ancestry male in 1KGP, who most likely did not have MMA cblB type, which is a neonatal-onset disorder that results in severe disability and sometimes death without treatment. A plausible explanation is that the three affected individuals from the literature happened to carry this putatively benign allele, and due to inadequate information about its frequency this allele was mistakenly associated with MMA. Since the P submission, GeneDx used variant classification criteria to interpret this variant as VUS, citing the relatively high variant frequency as evidence for benignity. Invitae (with criteria) and Natera (without criteria) have interpreted the variant as B.

We examined the ClinVar variant responsible for the single indicated affected individual in 2020 and found this variant could plausibly cause disease. GenMed Metabolism Lab submitted this variant, G50R in *OTC*, an X-linked gene, in 2014 and cited an article in which researchers found this variant in a male with late-onset Ornithine transcarbamylase deficiency (OTCD) but did not provide the age of onset^64^. OTCD is known to have a variable age of onset in a sizeable fraction of cases, and researchers have identified one individual who was 44 years old when disease onset began^42^. Plausibly, this variant may be associated with late onset OTCD and the 1KGP hemizygous South Asian ancestry male (NA21124) has not yet reached the age of onset.

### HGMD variant reclassification

In 2014, 16 Select HGMD variants contributed to an inferred pathogenic genotype in at least one 1KGP individual (Table S7). By December 2020, 8 of these variants were reclassified to DM?, and an additional 3 DM variants were cataloged that contributed to an inferred pathogenic genotype. In total, we observed 19 Select variants in an inferred pathogenic genotype, which were present in 11 genes: *OTC* (4), *PAH* (3), *ASS1* (2), *CBS* (2), *CPT2* (2), *ACAD8* (1), *ACADS* (1), *ACADVL* (1), *SLC22A5* (1), *SLC25A13* (1), and *TAZ* (1). We did not evaluate the Full HGMD variants in detail, but we do note that of the 32 DM and DM? variants that contributed to an inferred pathogenic genotype in 2014, none were reclassified to a non-disease-causing category by 2020.

HGMD rarely provides explanations for variant reclassification, so it is difficult to directly investigate why certain variants were reclassified. Instead, we examined the evidence for pathogenicity of the 19 Select variants identified in an inferred pathogenic genotype in 2014, 2016, or 2020. For each variant, we reviewed the articles cited by HGMD (Table S7). According to the cited articles, researchers observed these variants in probands who were diagnosed with an IEM. None of the articles provided evidence for pathogenicity equivalent to the ACMG/AMP guidelines, which is not surprising given that most of the articles were published prior to 2015. Additionally, 12 out of 19 studies (63%) did not show any direct evidence for the functional effect of the variant, such as experimental assays of gene expression or enzymatic activity, and therefore did not conclusively assign pathogenicity to the variant (Table S7). Assay absence was highly correlated with later reclassification from DM to DM?. Of the 5 variants classified as DM in 2014 for which assays were performed, all remained DM through 2020. Of the 11 variants for which no assay was performed, 8 were reclassified to DM? by 2020. Despite the predictive power of assay presence, the results of the assays were not always conclusive. For example, we found one 1KGP individual was homozygous for the variant c.1108A>G (M370V) in *ACADS*, which was cataloged by HGMD as DM (Table 2). Yet, the original article cited by HGMD indicates that the variant c.1108A>G has a much more mild effect on tetramerization than all other putatively pathogenic variants tested^65^. Similarly, functional assays of the variant c.1105C>T (R369C) in *CBS* in a yeast model indicated no effect on enzyme function in the article cited by HGMD^66^ (Table 2). Among the 19 studies cited by HGMD for these Select variants, only three studies directly measure the enzymatic activity of the observed variant^67-69^.

**Table 2:** Subset of HGMD variants seen in a pathogenic genotype in 1KGP. See Table S7 for full list of HGMD variants.

| Chr | Position | Ref | Alt | Gene | cDNA,<br>protein | # hom<br>or hemi<br>in 1KGP | # comp<br>het in<br>1KGP | 1KGP<br>sample<br>ID(s) | Variant-specific<br>assay | Outcome | PubMed<br>ID | HGMD<br>2014 | HGMD<br>2020 |
| --- | --- | --- | --- | --- | --- | --- | --- | --- | --- | --- | --- | --- | --- |
| 12 | 120739317 | A | G | ACADS | c.1108A>G<br>p.M370V | 1 | 0 | NA20878 | In vitro activity in<br>mouse<br>mitochondria | Very mild effect on<br>protein misfolding | 18523805 | DM | DM |
| 21 | 43060481 | G | A | CBS | c.1105C>T<br>p.R369C | 0 | 2 | HG02645<br>NA20289 | Yeast model | No effect on<br>enzyme function | 9361025 | DM | DM |
| X | 38381417 | C | T | OTC | c.374C>T<br>p.T125M | 1 | 0 | NA19117 | Biochemical test | OTC liver<br>activity < 1% | 8807340 | DM | DM |

One of these studies described the variant c.374C>T (T125M) in *OTC*, which was observed in a male newborn who died at the age of 14 days^67^. A biochemical assay verified the variant OTC enzymatic activity was <1% that of wild type in liver tissue. Surprisingly, we observed that one African ancestry male, NA19117, possesses c.374C>T in his single copy of the X-linked *OTC* gene. Although the genotype in this individual was called with low quality, this same genotype was re-confirmed with high genotype quality (GQ=187) by Complete Genomics (Table S3).

We observed a single variant (ACADS:c.1108A>G) that led to an inferred pathogenic genotype in 1KGP that was present in both the Select ClinVar and Select HGMD datasets. 6 variants that led to an inferred pathogenic genotype were shared by the Full ClinVar and Select HGMD datasets. A total of 8 variants that led to an inferred pathogenic genotype were shared by the Full ClinVar and Full HGMD datasets. By the end of 2020, 7 of these 8 variants were reclassified to a non-pathogenic category in ClinVar, while in HGMD, 4 of the variants were classified as DM, and 4 were classified as DM?

### Considering expanded stand-alone benign guidelines

We next considered how the use of updated BA1 guidelines changed the number of 1KGP individuals who were indicated affected. In accordance with the updated 2018 BA1 guidelines^45^, we removed variants from consideration that had a MAF > 5% in any gnomAD exomes continental population. This had no effect on our analysis of Select or Full ClinVar variants (Fig. S2A,B). Applying these guidelines to Select HGMD variants led to the removal of 1 variant in 2014 and 2016 (Table S5, Fig. S2C). We found that this reduced the Select HGMD indicated affected individuals by 15 African ancestry individuals in 2014 and 2016, while the 2020 individuals remained at 11. We next applied these guidelines to the Full HGMD variants, which led to the removal of 8 variants in all three years and reduced the number of indicated affected individuals by 75% in 2014 and 62% in 2020 (Fig. S2D). Additionally, there was no remaining significant ancestry skew after correcting for multiple tests (see Methods). When we implemented the 2018 BA1 guidelines, the ClinVar and HGMD datasets had similar rates of false positive individuals in 2014, and only recently have their rates diverged (Fig. S2E).

### Comparison of inferred incidence with known incidence of screened IEMs

Next, we sought to characterize the extent of misclassified rare variants that could not be removed by a MAF filter or identified as part of an inferred pathogenic genotype. To do this, we compared the screened IEM incidence inferred from each database with the known incidence of screened IEMs. The aggregate incidence of screened IEMs is estimated to be 1 in 3,200 births^52^. This includes a small number of X-linked IEMs, which are extremely rare, with an estimated aggregate incidence of 1 in 450,000 births. We used these values as baselines to compare with the inferred incidence of screened IEMs. We inferred the screened IEM incidence of each database from the 1KGP MAF of variants classified as pathogenic (see Methods), after applying the 2018 BA1 guidelines. Since the inferred incidence of X-linked IEMs is primarily determined by hemizygous males, we consider autosomal and X-linked IEMs separately. For autosomal IEMs, we found that both Full and Select ClinVar variants inferred an incidence greater than the known incidence prior to 2018 (Fig. S3A,C). By 2018, the inferred incidence fell below the known incidence, and has remained at 20% of the known incidence for both datasets. For X-linked IEMs, Select ClinVar variants have indicated an incidence of zero since 2014 (Fig. S3B). However, Full ClinVar variants have always suggested an incidence orders of magnitude higher than the known incidence, although since 2017 this has been due to just a single variant which primarily is found in East Asian ancestry (Fig. S3D). The more comprehensive perspective provided by screened IEM incidence also allows us to observe patterns that were too subtle to be seen in our analysis of indicated affected individuals. For example, we observed that a large fraction of the screened IEM incidence was skewed towards European ancestry from 2015 to 2017 in Select ClinVar variants (Fig. S3A). However, due to the extreme rarity of these conditions, it is difficult to precisely infer incidence from 1KGP.

When we considered Select HGMD autosomal variants, we found that the inferred screened IEM incidence has decreased slightly over time, yet in 2020 is triple the known incidence (Fig. S4A). The incidence inferred from Full HGMD autosomal variants has increased over time, and in 2020 was 10-fold greater than the known incidence (Fig. S4C). As with Full ClinVar variants, the X-linked IEM incidence suggested by Select and Full HGMD variants is orders of magnitude higher than the known incidence (Fig. S4B,D). The separation of autosomal and X-linked IEMs suggests that African ancestry skew remains among Full HGMD autosomal variants (Fig. S4C), but in our analysis of indicated affected individuals (Fig. S2D) this African ancestry skew is largely masked by X-linked variants with high MAF.

### Replication of major results

We replaced 1KGP with gnomAD 3.0 genomes to assess the reproducibility of our major findings. We considered gnomAD individuals from the five continental ancestries (African, Latino, East Asian, European, and South Asian; n = 63,269). gnomAD 3.0 does not include any individuals sampled in 1KGP. However, gnomAD does include individuals enrolled in genetic studies. Additionally, gnomAD does not provide individual-level data, so we were unable to identify compound heterozygous variants. These are significant limitations that restrict our confidence in absolute values derived from this analysis. Instead, we focus on robust claims that can be made from trends over time in the relative values we obtained.

For each cataloged variant, we recorded the number of homozygotes and hemizygotes in gnomAD. Overall, our gnomAD analysis replicated all major findings from our 1KGP analysis (Figs. S6-S10). Across both ClinVar and HGMD, we found that the proportion of individuals in gnomAD that were indicated affected was almost always less than the proportion indicated affected in 1KGP, but not by less than 50%. One exception was the number of gnomAD individuals indicated affected by Full and Select HGMD variants, which was one third of the size expected based on our 1KGP analysis (Fig. S6E,F).

The direction of change in indicated affected individuals for all four datasets over time was nearly always consistent with our 1KGP analysis. We found one notable difference when we considered indicated affected individuals using Select ClinVar variants. In 2015 and 2016, we found an unexpectedly large number of indicated affected individuals (Fig. S6C). This can be attributed to a single variant (ACADS:c.511C>T) with a gnomAD MAF >3% and which was P with 1 review star in 2015 and 2016. Due to its modest size, 1KGP did not contain any individuals indicated affected by this variant, although the existence of such individuals was suggested by our incidence analysis in 1KGP, which found elevated European ancestry incidence in Select ClinVar variants from 2015 to 2017 (Fig. S3A). The variant, which was classified as P with 1 star in 2015 and 2016, is more prevalent in European ancestry individuals, resulting in European ancestry individuals significantly (p < 3.8e×10^−10^) over-represented in 2015 and 2016 Select ClinVar variants, with an odds ratio of 4.0 (95% CI:2.4-6.7). When considering Full ClinVar variants, both East Asian (p < 4×10^−6^) and European (p < 1×10^−4^) ancestry individuals were significantly over-represented from 2014 through 2016, with East Asian individuals having an OR of 4.5 (95% CI:2.6-7.6). These results were obtained by applying the 2015 BA1 guidelines, and they were unaltered when the 2018 BA1 guidelines were applied.

In addition to confirming the African ancestry skew in indicated affected individuals in our 1KGP analysis of HGMD variants, we discovered significant ancestry skew (p < 2×10^−6^) towards East Asian ancestry individuals in Full HGMD variants in 2014 and 2016 (Fig. S6F), as well as significant skew towards European ancestry individuals (p < 2×10^−5^) in Select HGMD variants in 2020 (Fig. S6E). When 2018 BA1 guidelines were applied, significant skew remained for East Asian and European ancestry individuals (Fig. S7E,F). Due to the imbalanced ancestry composition of gnomAD, the described ancestry skew is not obvious from visual inspection of the figures.

With the greater number of individuals in gnomAD relative to 1KGP, we were able to directly compare the false positive rate of Select and Full ClinVar variants. Although there were fewer gnomAD individuals indicated affected by Select ClinVar variants, when considering the indicated affected individuals per cataloged variant, we found that there was little difference between Select and Full ClinVar variants.

### Comparison of variant reclassification between ClinVar and HGMD

Our analysis of reclassified variants has so far considered only those variants which contributed to an inferred pathogenic genotype in 1KGP individuals. To identify broad trends in variant reclassification in ClinVar and HGMD, we considered all screened IEM variants that were reclassified in ClinVar or HGMD between 2014 and 2020.

Out of 16,857 ClinVar variants, 3,772 (22%) were reclassified between April 2014 and December 2020 (Fig. S12). Of these reclassified variants, 28% were reclassified 2 or more times. To simplify our analysis, for each variant we considered only the variant’s classification when it first entered ClinVar and the variant’s classification at the end of 2020. Of the 4,917 P/LP variants in ClinVar between 2014 and 2020, we found 1,655 (34%) were reclassified by the end of 2020 (Fig. 2a). 78% of these reclassifications were towards greater evidence for pathogenicity, and the remaining 22% were towards reduced evidence for pathogenicity (8% of all P/LP variants). The most common reclassification towards greater evidence for pathogenicity was from P/LP 1 star to P/LP 2 stars. The most common reclassification towards reduced evidence for pathogenicity was from P/LP 1 star to Conflicting.

HGMD screened IEM variants were reclassified substantially less often than those in ClinVar. Out of 4,777 variants classified as DM or DM? in 2014 or 2016, just 40 (0.8%) were reclassified. 7 of these reclassifications were from DM? to DM, and the remaining 33 were towards reduced evidence for pathogenicity (0.7% of all DM or DM? variants). The most common reclassification towards reduced evidence for pathogenicity was from DM to DM?. Between 2014 and 2020, only 6 DM or DM? variants were retired.

When considering variants reclassified towards reduced evidence for pathogenicity, we found that ClinVar variants were reclassified at a rate 11-fold greater than those in HGMD. We recognize this analysis is impacted by the greater number of available time samples and variant categories in ClinVar compared to HGMD. However, when we repeat this analysis considering only ClinVar variants at time points for which HGMD data is available, while also collapsing ClinVar pathogenic variants to just 2 categories (see Methods), this result stands.

### Variant reclassification rates by ancestry

In our earlier analysis, we identified ancestry skew in likely misclassified variants. Next, we investigate whether ancestry influences overall reclassification rates of variants in ClinVar. Historically, large-scale exome and genome sequencing projects (from which MAF is often derived) have undersampled non-European individuals^70,71^. Thus, we suspected that non-European individuals may shoulder a larger burden of variants that were initially classified as P or LP due to uncertain MAF and later reclassified to be VUS or Conflicting. At the same time, we recognized that the largest ClinVar submitters are located in countries where a majority of the population has European ancestry. Consequently, variants common in European ancestry individuals may have a greater chance of being interpreted by multiple submitters which could lead to Conflicting interpretations.

To distinguish which of these effects likely dominate in ClinVar, we determined whether variants present in specific ancestries were disproportionately likely to be reclassified. First, for each variant we used gnomAD exomes to identify the continental ancestry group with the highest MAF, and we assigned the variant to that ancestry group. gnomAD exome MAFs were normalized to avoid bias from sample size differences between ancestries (see Methods). We first considered variants for which the classification was reduced in confidence, which includes P/LP and B/LB variants that were reclassified to VUS or Conflicting. For those variants that could be assigned to an ancestry, we visualized reclassifications using Sankey diagrams in which line width represents the number of reclassified variants, and lines were colored by ancestry (Fig. 2C). We observed that European ancestry variants were the largest group in most reclassification paths. However, this analysis did not account for the differences in ancestry composition of the variants submitted to ClinVar. To control for this potential bias, for each ancestry we normalized by both the number of variants assigned to that ancestry and the duration in which they were in ClinVar which we measure in variant-months (see Methods). One variant-month is equivalent to a single variant classified in ClinVar for one month. We normalized only by variants that could have contributed to the reclassification (in this case, P/LP and B/LB). Controlling for the ancestry composition of variants in ClinVar, we found that variants for which European ancestry individuals had the highest MAF were reclassified towards greater uncertainty at a rate of ∼0.8% per variant-month (Fig. 2D). This was approximately twice the rate of reclassification for variants for which African, East Asian, or Latino ancestries (all p < 8×10^−5^) had the highest MAF (Fig. 2D). This is consistent with our observation that amongst all variants classified in ClinVar, a larger fraction of European ancestry variants were classified as Conflicting (Fig. S11). South Asian variants were also found to have elevated reclassification towards greater uncertainty of approximately 0.6% per variant-month, significantly higher than East Asian or African variants (both p < 2×10^−4^).

We also considered variants for which classification increased in confidence, which includes VUS or Conflicting variants that were reclassified to P/LP or B/LB. After visualizing these reclassifications with Sankey plots, we observed that in many reclassification paths, European variants were not the largest group (Fig. 2E), in contrast with reclassification paths towards less confidence. Indeed, when we normalized by the ancestry composition of variants in ClinVar, we found no significant difference between variants most common in African, East Asian, European, or South Asian ancestry, each of which was reclassified at ∼0.3% per variant-month (Fig. 2F). The exception were variants most common in Latino ancestry, which were reclassified at ∼0.1% per variant-month.

## Discussion

Variant databases are under continuous development and growth^24,25^. Several studies have attempted to capture this progress at different snapshots in time, although these studies have generally looked at different database elements, making comparisons across time difficult^26-28,72^. Here, we investigated not a single point in time, but evaluated systematically the same disorders over 6 years across two different databases. In both databases, we observed a decrease over time in the number of 1KGP individuals indicated affected by an IEM. Based on the high temporal resolution the ClinVar archives afford, we can see this change was most pronounced in 2016 through 2018 after the establishment of the 2015 ACMG/AMP guidelines and coincident with allele frequency resources such as ExAC. We believe screened IEMs provide an informative lens that reveals broader database trends that may be representative of thousands of rare genetic disorders.

Perhaps our most striking finding is the large difference between the number of affected individuals indicated by HGMD and ClinVar in 2020. However, this difference is not entirely surprising. HGMD states that its curation policy is “to err on the side of inclusion and enter a variant into the database even if its pathological relevance may be questionable” and uses DM? classifications for this purpose as well as frequency flags in its online interface^73^. On the other hand, the clinicians and genetic testing laboratories that contribute to ClinVar are typically concerned with the immediate clinical implications of a variant. While they don’t want to pass over variants that could explain proband phenotypes, they are also loath to misinform patients or begin unnecessary interventions that may be irreversible. Thus, ClinVar contributors are invested in maintaining a database of pathogenic variants with minimal false negatives and false positives. An additional factor may be the increasing use of assertion criteria in variants contributed to ClinVar, which compels contributors to delineate the pieces of evidence leading to a classification. In contrast, many journals (from which HGMD curates variants) do not require these pieces of evidence. Therefore, this analysis should not be seen as a duel between two competing databases, but rather a quantitative comparison of the outcomes of two distinct variant cataloging methods. These distinct methods led to the 100-fold difference between the false positive rate of individuals indicated affected by Full ClinVar variants and Full HGMD variants, observed in both 1KGP (Fig. 1G) and gnomAD (Fig. S6G). While the Full HGMD rate (∼25 indicated affected individuals per 1,000 cataloged variants) is still relatively low, our analysis allows us to quantify this difference in specificity between the two databases. It is possible that a clinical analysis using HGMD, which includes a greater number of variants than ClinVar, would result in a higher sensitivity analysis, but we are not able to assess false negatives in this study. Understanding the differences between these databases may be valuable to not only clinical researchers, but also to non-domain experts such as computational researchers, who sometimes use HGMD and ClinVar interchangeably to develop variant interpretation methods^74^.

Due to founder mutations, individual IEM variants are often enriched in a single ancestry. However, when we consider the total burden of all screened IEMs, continental ancestry groups appear to be affected at similar rates^52^. We found that African ancestry individuals were disproportionately indicated affected in 2014 when HGMD Select variants were considered, but this skew was resolved by 2020. Yet, all of the DM variants causing the ancestry skew in 2014 were reclassified to DM?. Thus, when considering HGMD Full variants, we found that significant African ancestry skew remained. Encouragingly, when we applied the 2018 BA1 guidelines, we observed no significant ancestry skew among Full or Select HGMD variants. This suggests that much of the observed ancestry skew is due to population-specific common variants. This likely reflects the historical lack of African ancestry samples in large sequencing projects^75,76^. HGMD in particular may be susceptible to these factors, since it catalogs variants directly from publications, including older literature that was written when common variants in African ancestry individuals were poorly characterized. When older studies are given the same credence as recent ones, these disparities are more likely to be perpetuated.

In addition to the 2018 BA1 guidelines, Whiffin et al.^77^ have also proposed disorder-specific MAF thresholds which are supported by recent ACMG/AMP guideline specifications^78^. For example, under this system *PAH* variants would have a stand-alone benign MAF threshold of 1.5% assuming a maximum incidence of 1 in 5,000 births. However, we decided not to pursue Whiffin et al. thresholds due to the heterogeneity of our disorders and complications arising from incomplete penetrance in some disorders. Additionally, many screened IEMs are significantly more common in one ancestry group, due to founder effects, which makes it difficult to define thresholds.

Among ClinVar variants that were reclassified, very rarely did the initial submitter change their interpretation, and instead nearly all were reclassified due to conflicting interpretations that largely included assertion criteria. We carefully examined 10 ClinVar variants which previously contributed to an inferred pathogenic genotype but have since been re-classified to a non-pathogenic interpretation. Eight of these variants are currently interpreted as Conflicting. For many variants, this is an accurate descriptor and reflects enduring disagreement among submitters. However, for some variants this may be a byproduct of ClinVar’s definition of Conflicting. Specifically, if any P or VUS classification includes assertion criteria (one review star), then regardless of the number of B or LB classifications submitted, the record remains Conflicting until the P or VUS submitter changes or retracts their submission. If a submitter is no longer active, then an older submission becomes impossible to change. Although this system has advantages (historical knowledge is not lost), it may also impede the resolution of variants and indicate conflict when there is large consensus. For example, c.323G>T in *ASS1* is currently listed in ClinVar as Conflicting, yet it has the following interpretations with at least 1 review star: 1 B, 4 LB, and 1 VUS. The VUS interpretation is from 2017, while the 5 B/LB interpretations are more recent. Although researchers have found that older variant classifications tend to be less accurate, their influence persists^32^. This is even more true for HGMD, which predates ClinVar and thus also contains a large fraction of older classifications. Given the rapid increase in our ability to determine variant MAF and predict variant pathogenicity, even in the past 5 years, it may be reasonable to require ClinVar submitters to refresh older interpretations. Under such a system, submitters would need to confirm their interpretations after several years, or the interpretations would be deprecated. This would reduce the influence of ‘zombie’ interpretations that persist although their submitter is no longer active. Regardless of the exact strategy, methods to confirm the validity of older classifications will be valuable.

Careful analysis of ClinVar and HGMD variants that infer pathogenic genotypes in 1KGP revealed a few variants that do not fit the model of screened IEM variants, which typically result in severe, highly-penetrant disorders that begin in infancy or early childhood. For example, we discovered inferred pathogenic genotypes in 1KGP that included c.512C>G in ACAD8 (associated with asymptomatic disease), c.374C>T in *OTC* (our results suggest this variant has incomplete penetrance), and c.148G>A in *OTC* (observed in late onset disease). Asymptomatic IEMs occur when a proband does not have any noticeable signs of disease, but their metabolites reveal a disease phenotype. These variants are generally classified as disease-causing due to their potential to cause disease. However, as our analysis shows, some individuals will be predicted to have a disease, even though they may never develop symptoms (asymptomatic disease, incomplete penetrance) or symptoms may appear much later in life (late-onset). That we are identifying these variants (as well as variants in diseases known to be asymptomatic such as *ACADS* and *PRODH*) implies the variant databases may in fact be performing better than our analysis suggests. It is possible that some of these variants do cause symptomatic disease in some individuals but disease has not manifested in the 1KGP individuals. These variants may inhabit a gray zone between pathogenic and benign, and their existence bolsters existing appeals to reconsider the binary paradigm of pathogenic and benign classifications^26,79^. Our work shows that a feature such as optional flags on a ClinVar or HGMD record would be useful. Submitters or curators could then flag entries for various non-standard features for which there is evidence, such as asymptomatic disease, incomplete penetrance, or late-onset in a disease that is typically early onset. This information would then be readily available, without the need to search through supplemental materials of cited publications or detailed explanations provided by submitters. This would be a step towards a classification system that recognizes that variant pathogenicity is multi-faceted, and it would also enable greater interpretability of variant classification data for non-domain experts.

Our gnomAD analysis supported the major findings or our 1KGP analysis. However, we noted a persistent issue in which the number of indicated affected gnomAD individuals was proportionally about half of that expected from our 1KGP results. This is potentially explained by our inability to identify compound heterozygotes. In IEM cohorts, a majority of pathogenic genotypes are caused by compound heterozygous variants^80^. However, in our 1KGP analysis, compound heterozygotes rarely exceeded 20% of indicated affected individuals, possibly due to high-frequency false positives that contributed to a disproportionately large number of homozygotes. Alternatively, the reduction in gnomAD indicated affected individuals may be caused by the imbalanced ancestry composition of gnomAD, specifically the large fraction of European genomes (which had few indicated affected individuals in our 1KGP analysis) compared with the relative paucity of South Asian or East Asian genomes (which contributed to a large fraction of the indicated affected 1KGP individuals). Despite gnomAD’s imbalanced ancestry composition, its greater size did allow us to compare the false positive rate of Select and Full ClinVar, suggesting that the false positive rate was similar for both datasets. Additionally, our gnomAD analysis revealed ancestry skew towards East Asian and European individuals in both ClinVar and HGMD that could not be definitively detected by 1KGP.

This work has several limitations. Among rare diseases, the variants associated with screened IEMs are unusually well-curated thanks to newborn screening programs. Thus, screened IEMs are not necessarily representative of many rare diseases. Furthermore, our primary analysis was limited by the comparably small size of 1KGP relative to the rarity of IEMs. At the same time, 1KGP has several advantages, including its approximately even representation of the 5 major continental ancestries and its open availability of genomes, which allowed us to identify individuals who are compound heterozygous for variants classified as pathogenic and to validate the quality of nearly all analyzed variants. These unique features give 1KGP enduring value. Our analysis was particularly sensitive to putatively misclassified variants on the X chromosome since we considered males who were hemizygous for an variant classified as pathogenic to be indicated affected. This explains the relatively high number of observed *OTC* and *TAZ* variants flagged by our analyses of ClinVar and HGMD, despite the extreme rarity of their associated disorders. Finally, since few ClinVar submitters provide detailed explanation for their interpretation, and HGMD does not provide detailed explanation for its classifications, for many variants it is difficult to determine with confidence why interpretations changed over time.

We have investigated how the false positive rate of ClinVar and HGMD variants has changed over time. Our results suggest that ClinVar has a lower false positive rate than HGMD due to variant reclassification occurring in the past few years. We noted patterns in variant reclassification and found that variant interpretation guidelines and diverse allele frequency databases principally contributed to these reclassifications. In agreement with the lower false positive rate of ClinVar variants, we found that variants classified as pathogenic in ClinVar are reclassified 11-fold more often than those in HGMD, suggesting that misclassified variants are more readily reclassified in ClinVar than HGMD. We also discovered that variants common in European and South Asian individuals were significantly more likely to be reclassified from P/LP or B/LB to VUS or Conflicting. We conclude that although the allele frequency of variants common in European individuals has been known for longer, due to the increased chance they will be classified by multiple submitters, they are more often reclassified from a confident category to a less confident category in ClinVar. We anticipate that this work will be a valuable benchmark of the progress that has been made in variant interpretation, of interest to the individuals who maintain these databases, the clinical researchers who use these databases regularly, and the computational researchers who use these databases for training and testing methods.

## Data Availability

All data produced are available online at https://doi.org/10.6078/D1872X

https://ftp.ncbi.nlm.nih.gov/pub/clinvar/vcf_GRCh38/

https://www.hgmd.cf.ac.uk/ac/index.php

https://www.internationalgenome.org/data

## Acknowledgements

We thank Zhiqiang Hu for valuable advice throughout the project. We thank Priya Moorjani, Dan Rokhsar, and Nilah Ioannidis for valuable comments. We thank members of the Brenner lab for their suggestions to improve the project. S.E.B. was unable to fully review this manuscript due to injury. A.G.S. was supported by a NSF Graduate Research Fellowship, Grant No. DGE 1752814, and the NSF Postdoctoral Research Fellowships in Biology Program under Grant No. 2109912. This work was also supported by NIH grant P01 AI138962, NIH grant U19 HD077627, NIH grant U41 HG007346, a research agreement with Tata Consultancy Services, and the Chan Zuckerberg Biohub.

