## Supplemental Figures for "ClinVar and HGMD genomic variant classification accuracy has improved over time, as measured by implied disease burden"

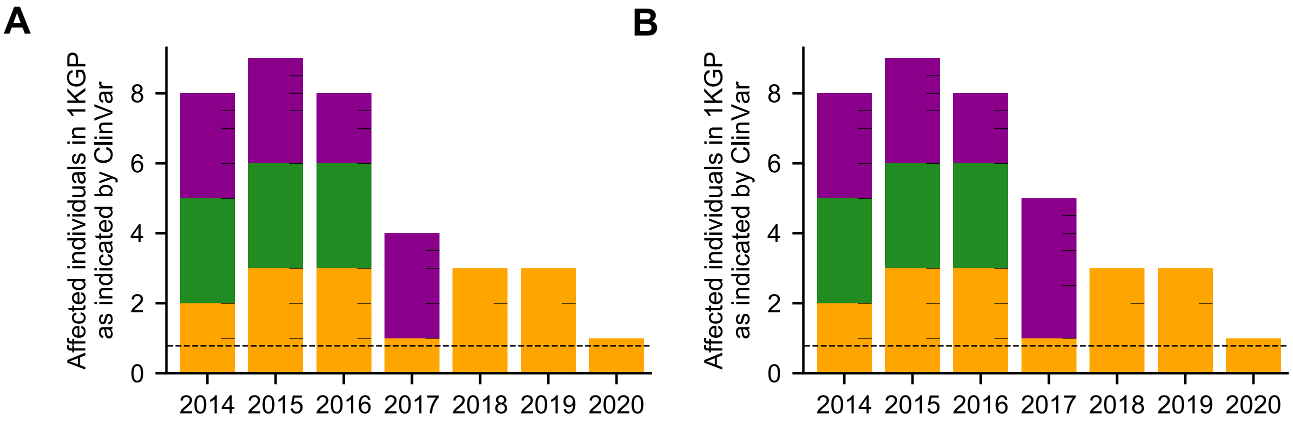


**Fig. S1.** (A) Indicated affected individuals in 1KGP when P variants with any number of stars are considered. (B) Indicated affected individuals in 1KGP when P or LP variants with any number of stars are considered (identical to Fig. 1D). The only difference is a single individual in 2017. Bar color, tick marks, and dashed lines are used as described in Fig. 1.

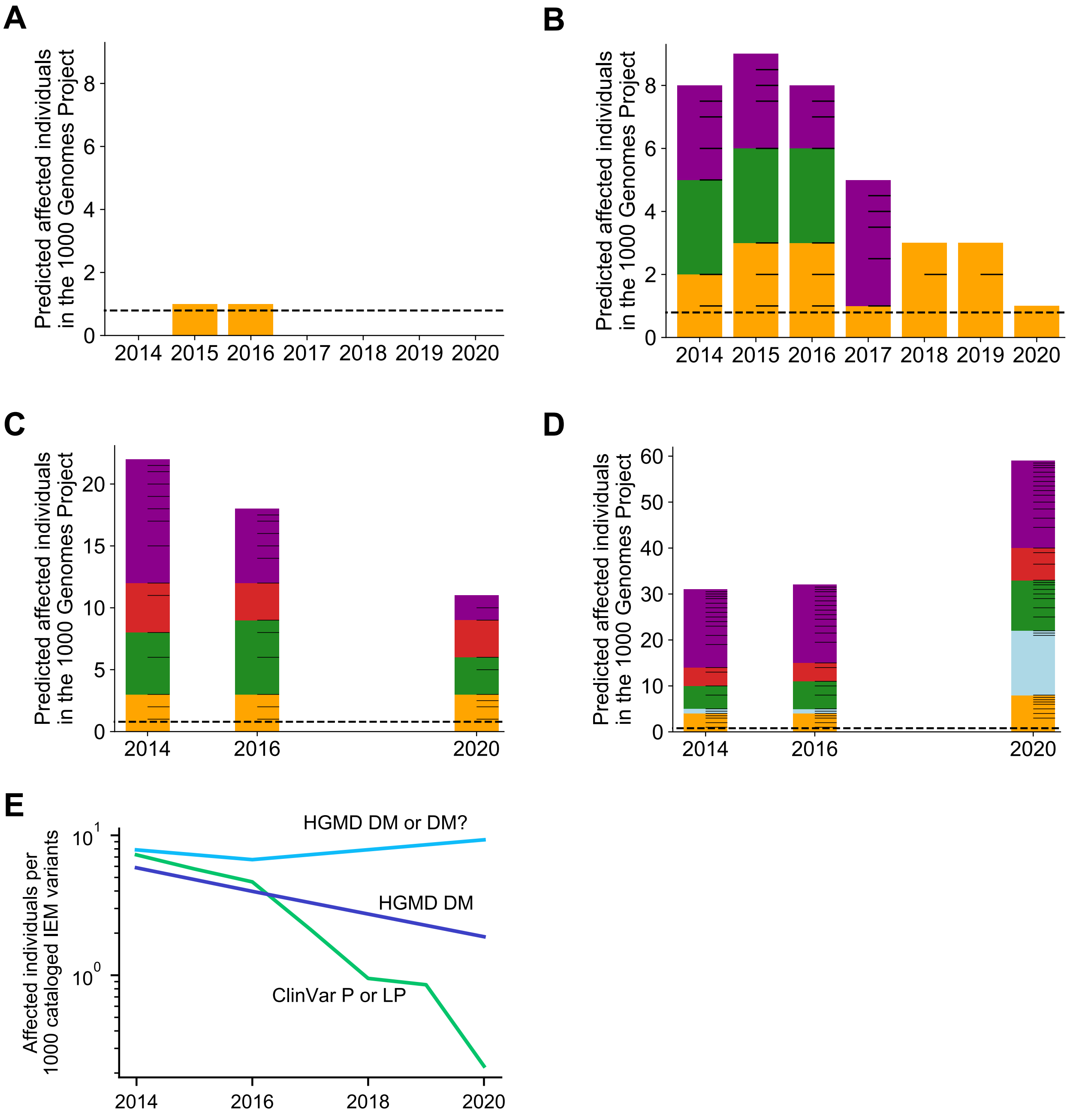


**Fig. S2.** Number of 1KGP individuals indicated affected for screened IEMs with the 2018 BA1 guidelines implemented. Bar coloring, tick marks, and dashed lines are used as described in Fig. 1. The number of 1KGP individuals with an implied pathogenic genotype for a variant in (A) Select ClinVar variants annotated as pathogenic. (B) Full ClinVar variants annotated as pathogenic. (C) Select HGMD variants. (D) Full HGMD variants. (E) The number of affected individuals relative to the number of variants classified in each variant set.

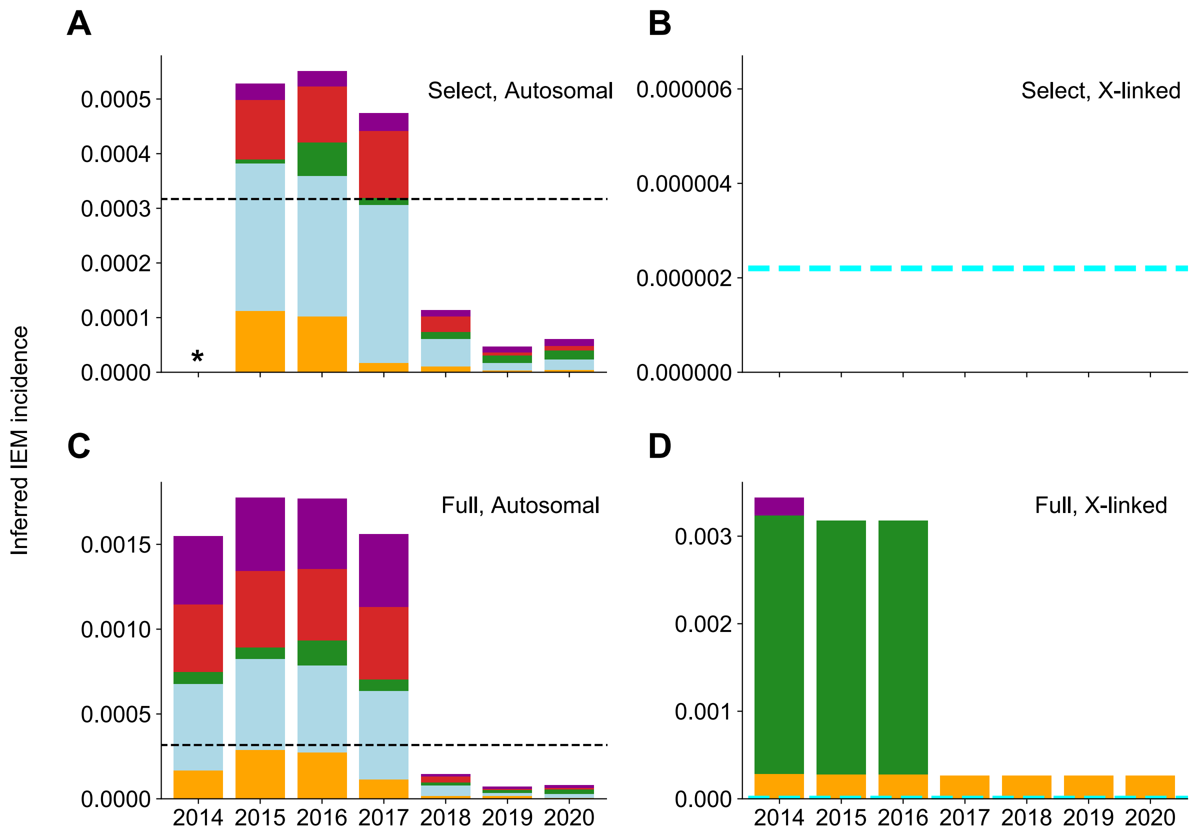


**Fig. S3.** Incidence of screened IEMs inferred by ClinVar variants. Dashed black line indicates expected incidence of autosomal screened IEMs of 1 in 3,200 births. Dashed cyan line indicates expected incidence of X-linked screened IEMs of 1 in 450,000 births. Bar colors are used as described in Fig. 4.1. The screened IEM incidence in 1KGP inferred by allele frequency of (A) Select autosomal ClinVar variants annotated as pathogenic. (B) Select X-linked ClinVar variants annotated as pathogenic. (C) Full autosomal ClinVar variants annotated as pathogenic. (D) Full X-linked ClinVar variants annotated as pathogenic. The 2015 BA1 guidelines were applied. When the 2018 BA1 guidelines are applied, the results did not change. *Data not available because existing review star framework was not in place until 2015.

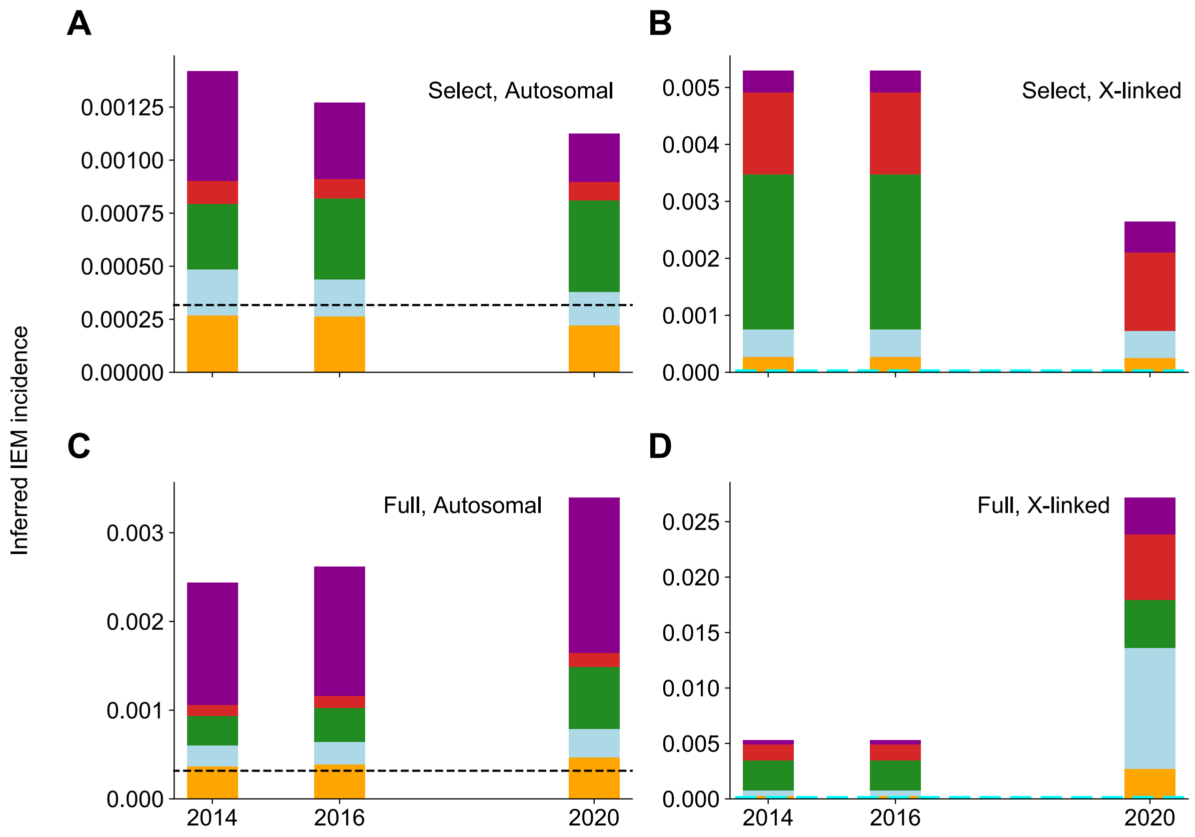


**Fig. S4.** Incidence of screened IEMs inferred by HGMD variants. Dashed black line indicates expected incidence of autosomal screened IEMs of 1 in 3,200 births. Dashed cyan line indicates expected incidence of X-linked screened IEMs of 1 in 450,000 births. Bar colors are used as described in Fig. 4.1. The screened IEM incidence in 1KGP inferred by allele frequency of (A) Select autosomal HGMD variants. (B) Select X-linked HGMD variants. (C) Full autosomal HGMD variants. (D) Full X-linked HGMD variants. The 2018 BA1 guidelines were applied.

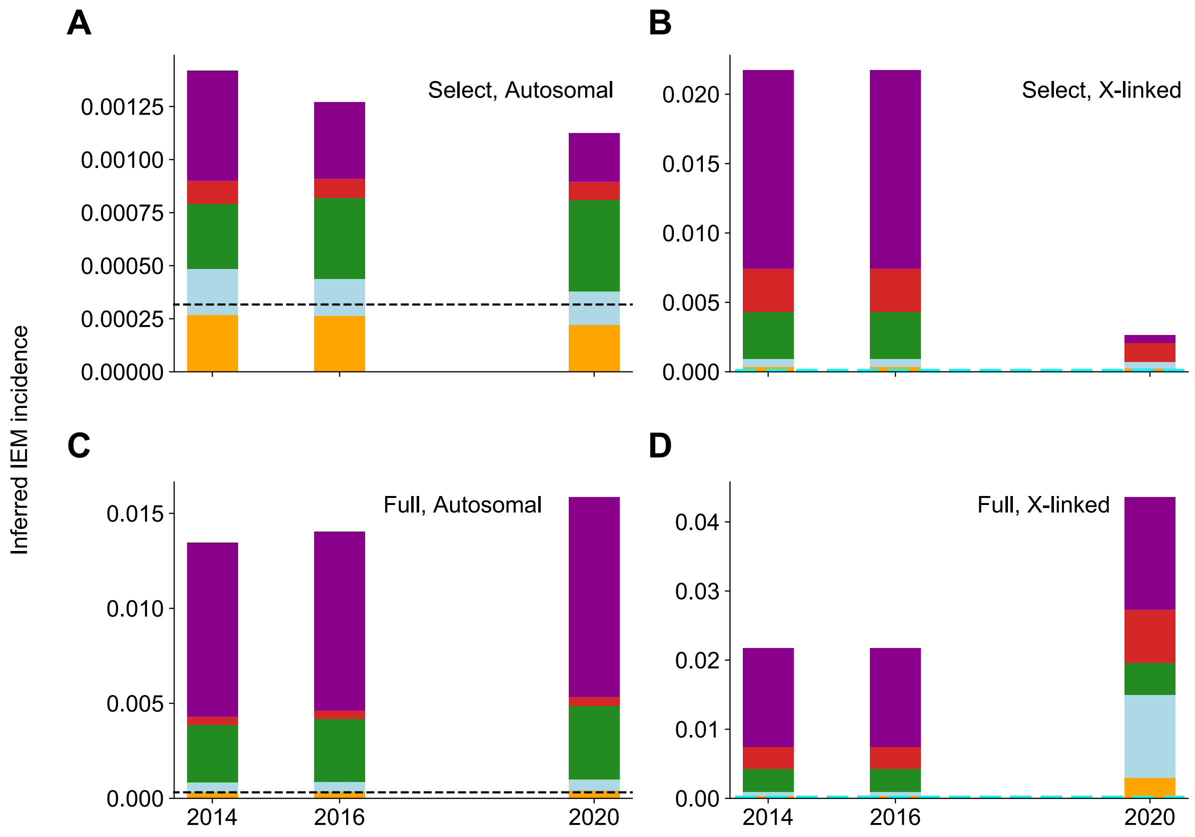


**Fig. S5.** Incidence of screened IEMs inferred by HGMD variants. Dashed black line indicates expected incidence of autosomal screened IEMs of 1 in 3,200 births. Dashed cyan line indicates expected incidence of X-linked screened IEMs of 1 in 450,000 births. Bar colors are used as described in Fig. 4.1. The screened IEM incidence in 1KGP inferred by allele frequency of (A) Select autosomal HGMD variants. (B) Select X-linked HGMD variants. (C) Full autosomal HGMD variants. (D) Full X-linked HGMD variants. The 2018 BA1 guidelines were applied.

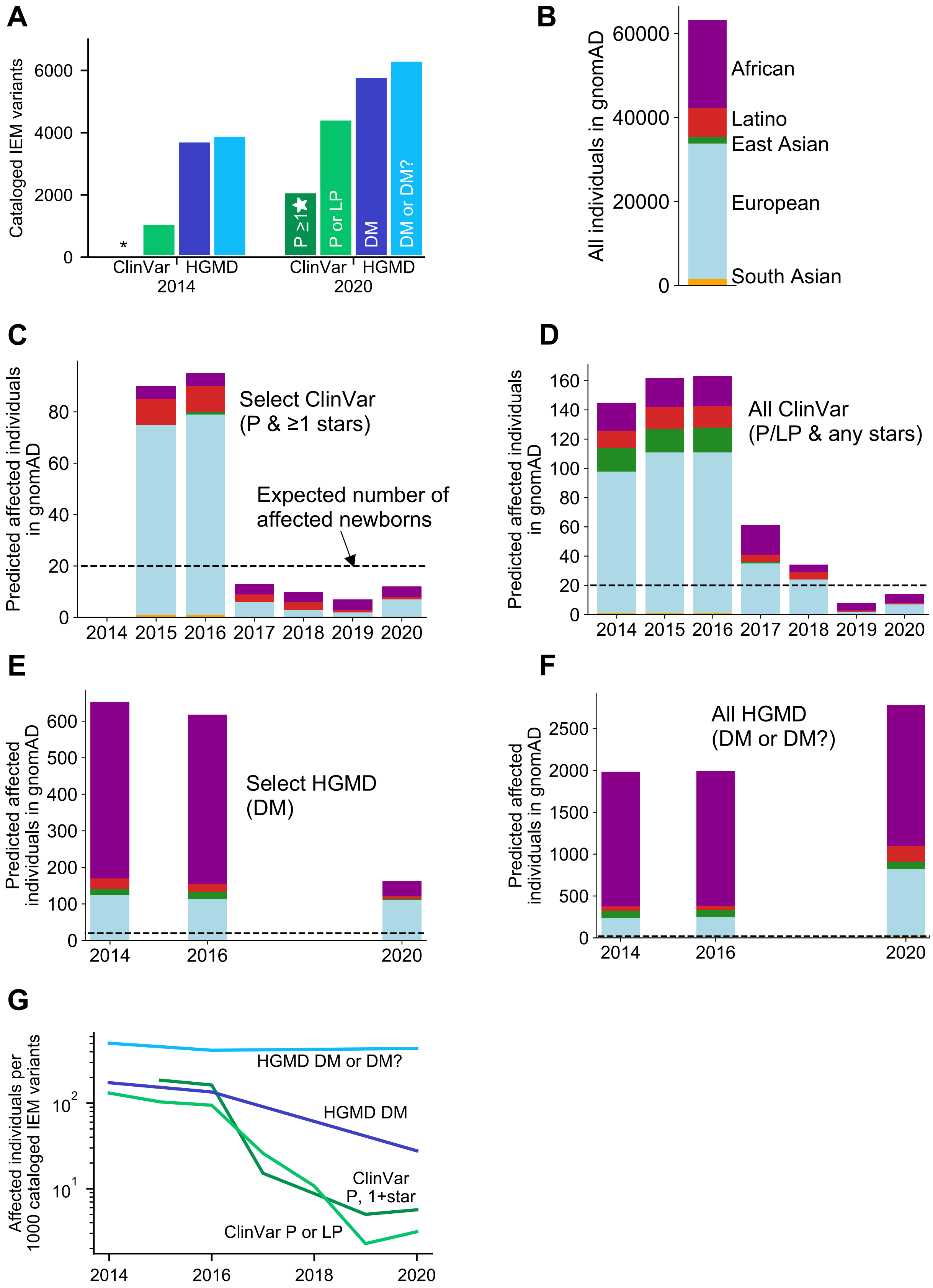
A
Cataloged IEM variants
6000
4000
2000
0
*
P ≥1
P or LP
DM
DM or DM?
ClinVar
HGMD
2014
ClinVar
HGMD
2020
B
All individuals in gnomAD
60000
40000
20000
0
African
Latino
East Asian
European
South Asian
C
Predicted affected individuals in gnomAD
Select ClinVar
(P & ≥1 stars)
Expected number of affected newborns
2014 2015 2016 2017 2018 2019 2020
D
Predicted affected individuals in gnomAD
All ClinVar
(P/LP & any stars)
2014 2015 2016 2017 2018 2019 2020
E
Predicted affected individuals in gnomAD
Select HGMD
(DM)
2014 2016 2020
F
Predicted affected individuals in gnomAD
All HGMD
(DM or DM?)
2014 2016 2020
G
Affected individuals per 1000 cataloged IEM variants
HGMD DM or DM?
HGMD DM
ClinVar P, 1+star
ClinVar P or LP
2014 2016 2018 2020

**Fig. S6.** Number of gnomAD individuals indicated affected for screened IEMs by ClinVar or HGMD over time. (A) Number of screened IEM variants present in ClinVar or HGMD in 2014 and 2020 (identical to Fig. 1A). (B) Ancestry composition of individuals in gnomAD. Due to the imbalanced ancestry composition, ancestry skew is difficult to discern visually in the following panels. (C-F) Bars are colored by ancestry as shown in (B). Dashed black lines indicate the aggregate population incidence of screened IEMs. The 2015 BA1 guidelines were applied. The number of gnomAD individuals with a pathogenic genotype for a variant in (C) Select ClinVar variants annotated as pathogenic, defined as variants with a P interpretation with at least 1 review star. Variants that also have conflicting interpretations (with VUS or B/LB) with 1 or more review stars are removed. (D) Full ClinVar variants annotated as pathogenic, defined as variants with a P or LP interpretation. Variants that also have conflicting interpretations (with VUS or B/LB) are removed. (E) Select HGMD variants, defined as variants classified as DM. 2014, 2016, and 2020 are shown because they are the years for which we have archived HGMD data. (F) Full HGMD variants, defined as variants classified as DM or DM?. (G) The number of affected individuals relative to the number of variants classified in each variant set. *Data not available because existing review star framework was not in place until 2015.

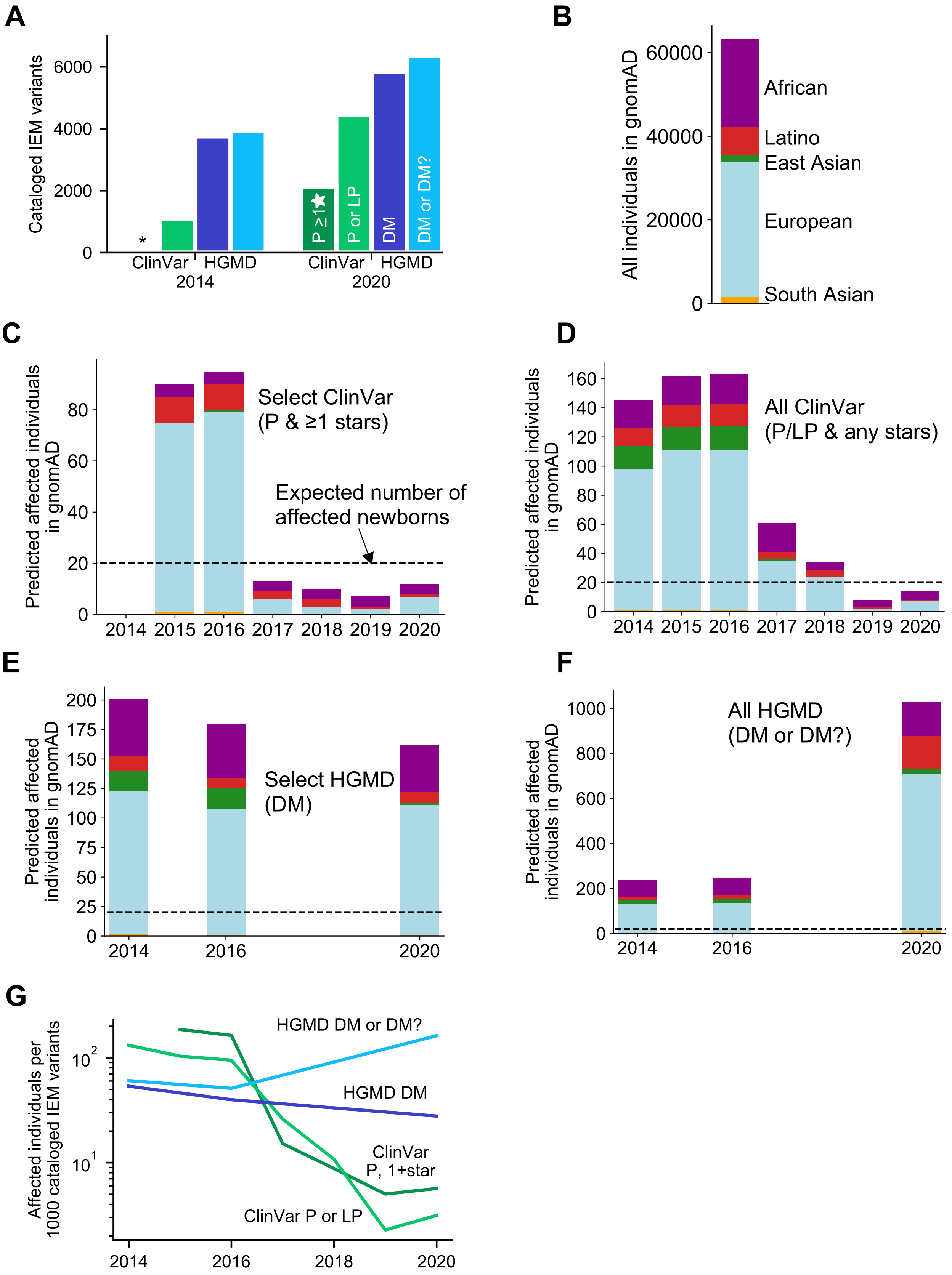
A
Cataloged IEM variants
6000
4000
2000
0
*
P ≥1★
P or LP
DM
DM or DM?
ClinVar
HGMD
2014
ClinVar
HGMD
2020
B
All individuals in gnomAD
60000
40000
20000
0
African
Latino
East Asian
European
South Asian
C
Predicted affected individuals in gnomAD
80
60
40
20
0
Select ClinVar
(P & ≥1 stars)
Expected number of
affected newborns
2014 2015 2016 2017 2018 2019 2020
D
Predicted affected individuals in gnomAD
160
140
120
100
80
60
40
20
0
All ClinVar
(P/LP & any stars)
2014 2015 2016 2017 2018 2019 2020
E
Predicted affected individuals in gnomAD
200
175
150
125
100
75
50
25
0
Select HGMD
(DM)
2014
2016
2020
F
Predicted affected individuals in gnomAD
1000
800
600
400
200
0
All HGMD
(DM or DM?)
2014
2016
2020
G
Affected individuals per 1000 cataloged IEM variants
$10^2$
$10^1$
HGMD DM or DM?
HGMD DM
ClinVar
P, 1+star
ClinVar P or LP
2014
2016
2018
2020

**Fig. S7.** Number of gnomAD individuals indicated affected for screened IEMs by ClinVar or HGMD over time. (A) Number of screened IEM variants present in ClinVar or HGMD in 2014 and 2020 (identical to Fig. 1A). (B) Ancestry composition of individuals in gnomAD. Due to the imbalanced ancestry composition, ancestry skew is difficult to discern visually in the following panels. (C-F) Bars are colored by ancestry as shown in (B). Dashed black lines indicate the aggregate population incidence of screened IEMs. The 2018 BA1 guidelines were applied. The number of gnomAD individuals with a pathogenic genotype for a variant in (C) Select ClinVar variants annotated as pathogenic, defined as variants with a P interpretation with at least 1 review star. Variants that also have conflicting interpretations (with VUS or B/LB) with 1 or more review stars are removed. (D) Full ClinVar variants annotated as pathogenic, defined as variants with a P or LP interpretation. Variants that also have conflicting interpretations (with VUS or B/LB) are removed. (E) Select HGMD variants, defined as variants classified as DM. 2014, 2016, and 2020 are shown because they are the years for which we have archived HGMD data. (F) Full HGMD variants, defined as variants classified as DM or DM?. (G) The number of affected individuals relative to the number of variants classified in each variant set. *Data not available because existing review star framework was not in place until 2015.

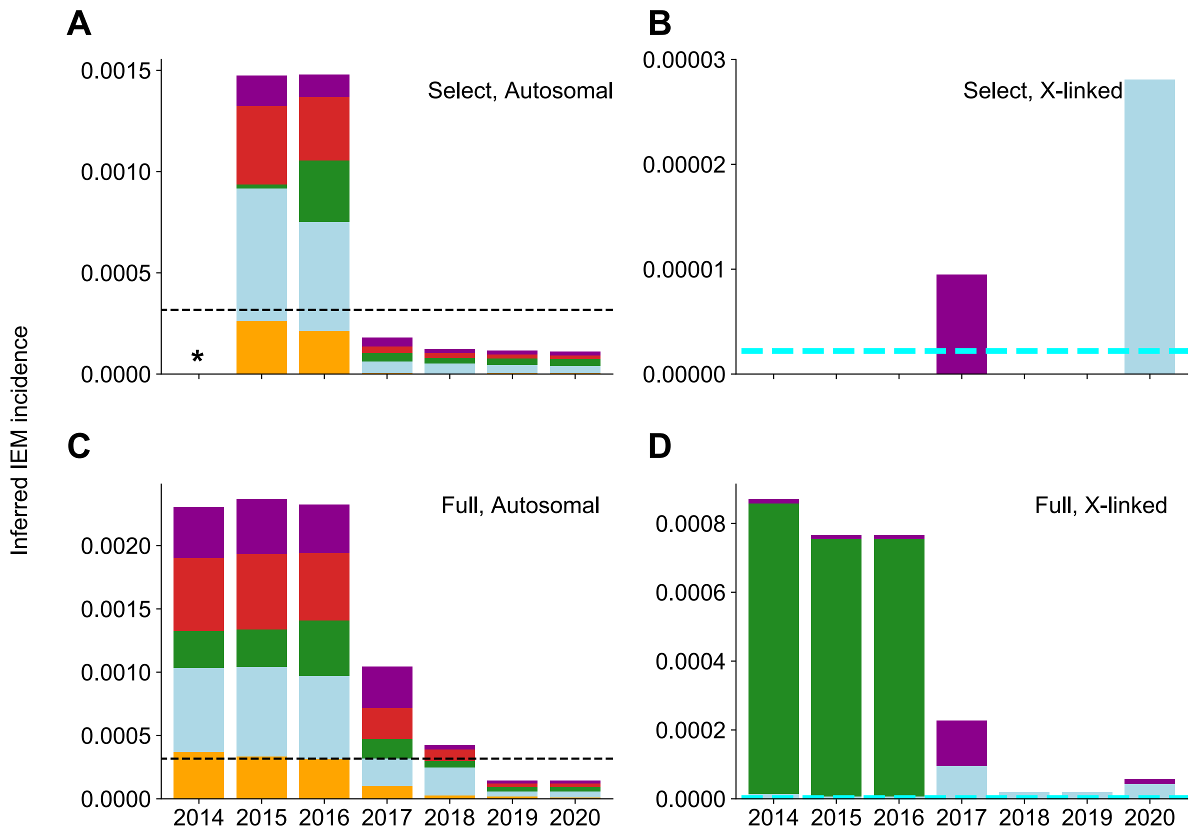


**Fig. S8.** Incidence of screened IEMs inferred from ClinVar variants and gnomAD allele frequencies. Dashed black line indicates expected incidence of autosomal screened IEMs of 1 in 3,200 births. Dashed cyan line indicates expected incidence of X-linked screened IEMs of 1 in 450,000 births. Bar colors are used as described in Fig. 4.1. The screened IEM incidence in gnomAD inferred by allele frequency of (A) Select autosomal ClinVar variants annotated as pathogenic. (B) Select X-linked ClinVar variants annotated as pathogenic. (C) Full autosomal ClinVar variants annotated as pathogenic. (D) Full X-linked ClinVar variants annotated as pathogenic. The 2015 BA1 guidelines were applied. When the 2018 BA1 guidelines are applied, the results did not change. *Data not available because existing review star framework was not in place until 2015.

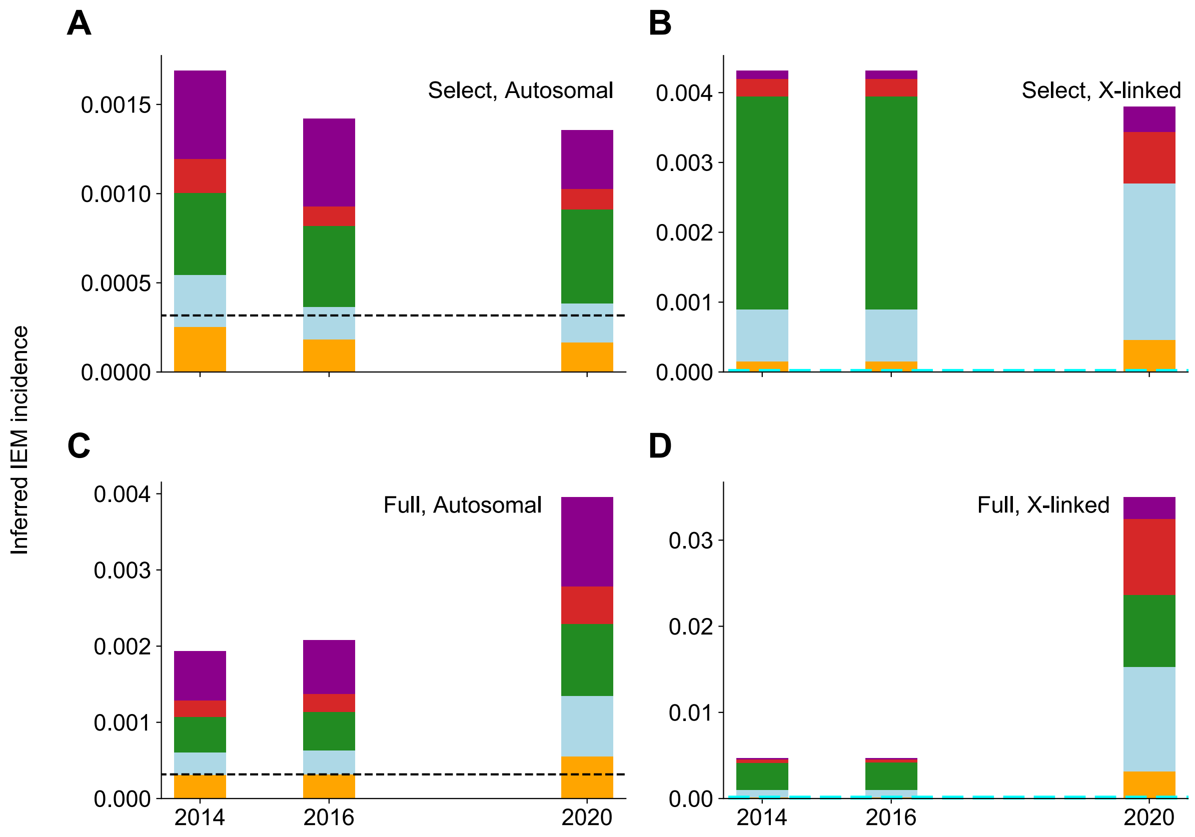


**Fig. S9.** Incidence of screened IEMs inferred by HGMD variants and gnomAD allele frequencies. Dashed black line indicates expected incidence of autosomal screened IEMs of 1 in 3,200 births. Dashed cyan line indicates expected incidence of X-linked screened IEMs of 1 in 450,000 births. Bar colors are used as described in Fig. 4.1. The screened IEM incidence in 1KGP inferred by allele frequency of (A) Select autosomal HGMD variants. (B) Select X-linked HGMD variants. (C) Full autosomal HGMD variants. (D) Full X-linked HGMD variants. The 2018 BA1 guidelines were applied.

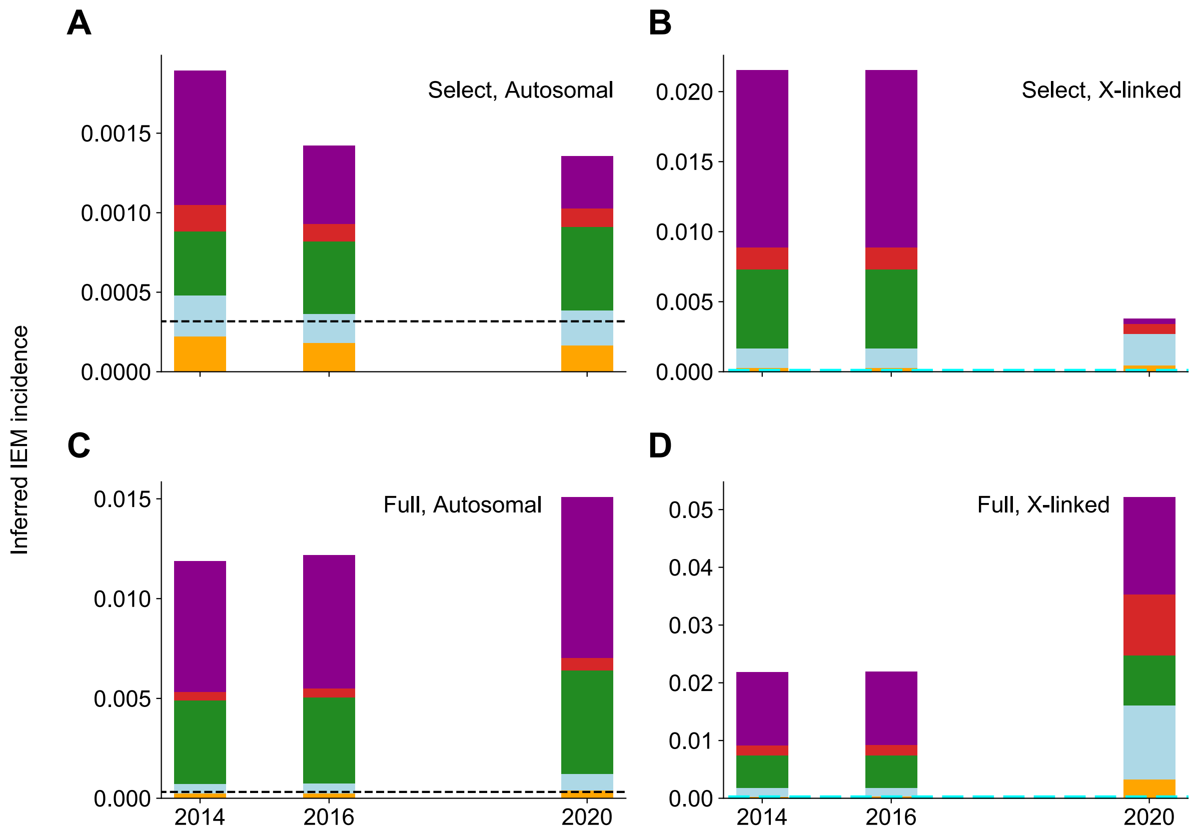


**Fig. S10.** Incidence of screened IEMs inferred by HGMD variants. Dashed black line indicates expected incidence of autosomal screened IEMs of 1 in 3,200 births. Dashed cyan line indicates expected incidence of X-linked screened IEMs of 1 in 450,000 births. Bar colors are used as described in Fig. 4.1. The screened IEM incidence in 1KGP inferred by allele frequency of (A) Select autosomal HGMD variants. (B) Select X-linked HGMD variants. (C) Full autosomal HGMD variants. (D) Full X-linked HGMD variants. The 2015 BA1 guidelines were applied.

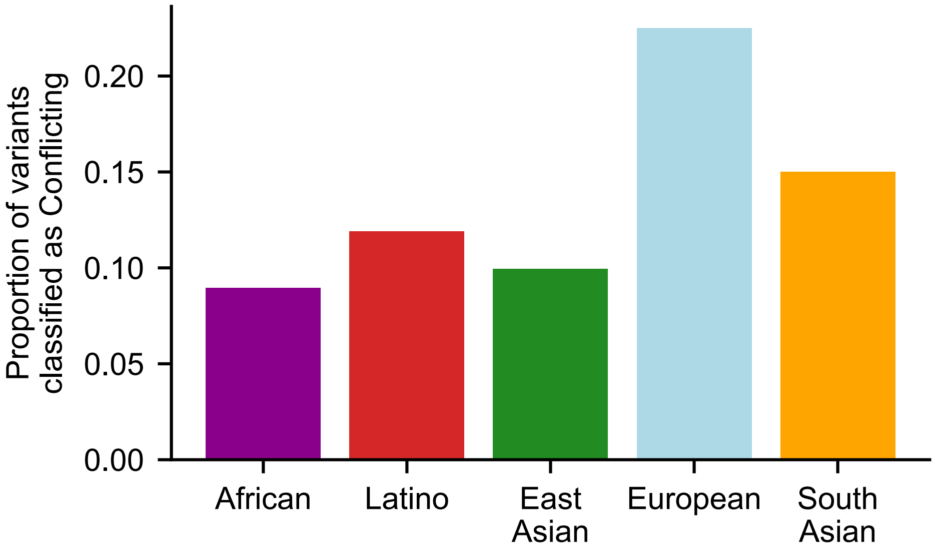


**Fig. S11.** For each ancestry, proportion of ClinVar variants that are classified as Conflicting. For each variant, we used gnomAD exomes to identify the ancestry group with the highest MAF, and we assigned the variant to that ancestry group.

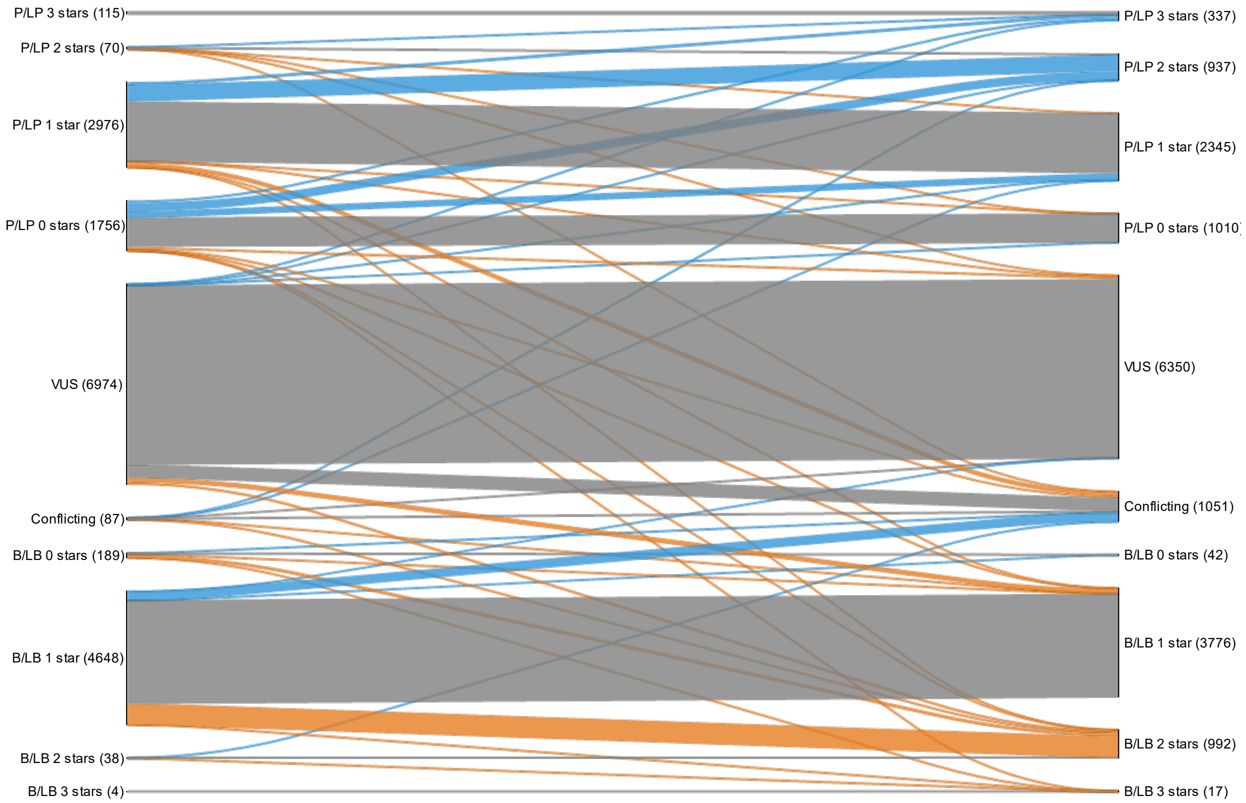


**Fig. S12.** Reclassification paths of all screened IEM ClinVar variants from 2014 (or first submission thereafter) to 2020, visualized in a Sankey plot in which line width represents the number of reclassified variants. Blue lines indicate increasing annotated pathogenicity or review stars, orange lines indicate increasing annotated benignity or reduced confidence of pathogenicity, and gray lines indicate no change. Numbers in parentheses provide variant counts of initial and final classifications for each category.
