## Supplemental Table 6 for "ClinVar and HGMD genomic variant classification accuracy has improved over time, as measured by implied disease burden"

| Chr | Position | Ref | Alt | Gene | cDNA, protein | Variant type | # hom or hemi in 1KGP | # comp het in 1KGP | 1KGP sample ID(s) | First pathogenic submission: Submitter, Date, Interpretation, Evidence | First non-pathogenic submission: Submitter, Date, Interpretation, Evidence | Consensus Interpretation as of Dec 2020* | Submitted Interpretations |
|---|---|---|---|---|---|---|---|---|---|---|---|---|---|
| 5 | 132384290 | C | T | SLC22A5 | c.713C>T<br>p.A238G | missense | 1 | 0 | HG04075 | Research and Development (ARUP Laboratories),<br>April 2014,<br>Pathogenic,<br>Present in affected individuals | EGL Genetic Diagnostics,<br>January 2017,<br>VUS,<br>Clinical testing | Conflicting Interpretations of Pathogenicity | 1 Pathogenic<br>3 Likely pathogenic<br>1 VUS |
| 6 | 49456101 | G | A | MMUT | c.890C>T<br>p.T297I | missense | 2 | 0 | HG03598<br>HG03686 | Institute of Medical Genetics and Genomics (Sir Ganga Ram Hospital),<br>October 2018,<br>Pathogenic,<br>None | Invitae,<br>December 2019,<br>Likely benign,<br>Clinical testing | Conflicting Interpretations of Pathogenicity | 1 Pathogenic NC<br>1 VUS,<br>1 Likely benign<br>1 Benign NC |
| 9 | 130452225 | C | T | ASS1 | c.-4=<br>NA | 5' UTR | 0 | 1 | NA19030 | GeneDx,<br>August 2015,<br>Pathogenic,<br>clinical testing | Invitae,<br>January 2017,<br>VUS,<br>Non-coding variant seen in affected and in population databases | Conflicting Interpretations of Pathogenicity | 2 Likely pathogenic<br>2 VUS |
| 9 | 130458549 | G | T | ASS1 | c.323G>T<br>R108L | missense | 1 | 1 | NA19030<br>NA19395 | OMIM,<br>April 2014,<br>Pathogenic,<br>Heterozygous variant in affected individual | Illumina,<br>April 2017,<br>VUS,<br>Observed in healthy population | Conflicting Interpretations of Pathogenicity | 1 Pathogenic NC<br>1 VUS<br>4 Likely benign<br>1 Benign<br>1 Benign NC |
| 12 | 109561798 | C | T | MMAB | c.403G>A<br>p.A135T | missense | 1 | 0 | HG03169 | GeneReviews,<br>February 2016,<br>Pathogenic,<br>Seen in affected individuals | GeneDx,<br>May 2017,<br>VUS,<br>Variant is conserved and predicted damaging, but seen at high frequency in African ancestry populations. | Conflicting Interpretations of Pathogenicity | 1 Pathogenic NC<br>1 VUS<br>1 Benign<br>1 Benign NC |
| 12 | 120739317 | A | G | ACADS | c.1108A>G<br>p.M370V | missense | 1 | 0 | NA20878 | GeneDx,<br>August 2015,<br>Pathogenic,<br>clinical testing | Illumina,<br>January 2017,<br>VUS,<br>Observed in both healthy and affected individuals, with high population allele frequency | Conflicting Interpretations of Pathogenicity | 2 VUS<br>1 Likely Benign |
| 22 | 18918386 | G | A | PRODH | c.1357C>T<br>p.R453C | missense | 0 | 1 | NA19372 | OMIM,<br>April 2014,<br>Pathogenic,<br>In vitro assay | Laboratory for Molecular Medicine (Partners HealthCare Personalized Medicine),<br>May 2017,<br>Benign,<br>Elevated population allele frequency | Conflicting Interpretations of Pathogenicity | 1 Pathogenic NC<br>1 VUS<br>1 Likely benign<br>1 Benign |
| 22 | 18918421 | A | G | PRODH | c.1322T>C<br>p.L441P | missense | 0 | 1 | NA19372 | OMIM,<br>April 2014,<br>Pathogenic,<br>In vitro assay | Invitae,<br>December 2017,<br>VUS,<br>Highly conserved, with in vitro evidence, but high allele frequency | Conflicting Interpretations of Pathogenicity | 1 Pathogenic NC<br>3 Likely pathogenic<br>1 VUS |
| X | 38367361 | G | A | OTC | c.148G>A<br>p.G50R | missense | 1 | 0 | NA21124 | GenMed Metabolism Lab,<br>April 2014,<br>Pathogenic,<br>Identified in late onset individual | None | Pathogenic, 0 stars | 2 Pathogenic NC |
| X | 38369882 | G | C | OTC | c.298+5G>C<br>NA | Donor splice site | 3 | 0 | HG00622<br>HG01844<br>HG02073 | GenMed Metabolism Lab,<br>April 2014,<br>Pathogenic,<br>Identified in affected female | EGL Genetic Diagnostics,<br>January 2017,<br>Benign<br>Classified in clinical testing | Benign/Likely benign | 1 Pathogenic NC<br>1 VUS NC<br>1 Likely benign<br>5 Benign<br>1 Benign NC |
| X | 38381417 | C | T | OTC | c.374C>T<br>p.T125M | missense | 1 | 0 | NA19117 | GenMed Metabolism Lab,<br>April 2014,<br>Pathogenic,<br>Identified in affected individual | University of Washington,<br>January 2015,<br>VUS,<br>Classified as part of investigation of incidental findings in population cohort | VUS | 3 VUS<br>1 Pathogenic NC |

Table S6

**Table S6.** ClinVar variants found in a pathogenic genotype in one or more predicted affected individuals from 1KGP. *ClinVar only considers variants with criteria in determining consensus interpretation. NC = No assertion criteria provided.

I note that for the X chr genes, not sure that 'Hom" is the best descriptor. Maybe that column should be Hom/Hemi?

The exact transcript versions that I used for the HGVS in the above genes

SLC22A5: NM_001308122.2, NP_001295051.1

MMUT: NM_000255.4, NP_000246.2

ASS1: NM_000050.4, NP_000041.2

MMAB: NM_052845.4, NP_443077.1

ACADS: NM_000017.4, NP_000008.1

PRODH: NM_016335.6, NP_057419.5

OTC: NM_000531.6, NP_000522.3
